# CHI3L1 (YKL-40) and Chit-1 expressing glia in the white matter of ALS, FTLD, and AD correlate to pathology and disease duration

**DOI:** 10.1101/2025.05.05.25326702

**Authors:** Chelsea Tran, Nisha Reddy, J.K. Thomas, Vinisha Venugopal, Robert Bowser

## Abstract

**Background:** Chitotriosidase (Chit-1) and chitinase-3-like protein 1 (CHI3L1) protein levels are increased in the cerebrospinal fluid (CSF) of neurodegenerative diseases, including amyotrophic lateral sclerosis (ALS), frontotemporal dementia (FTD), and Alzheimer’s disease (AD). Few studies have examined the spatial expression of chitinase expressing cells with respect to neuropathologic hallmarks of disease.

**Methods:** RNA-sequencing was used to examine Chit-1 and CHI3L1 gene expression in the spinal cord and motor cortex. Immunohistochemistry was used to characterize the distribution of Chit-1 and CHI3L1 expressing cells in ALS, C9-ALS, FTLD, AD, and non-neurologic disease controls. Immunofluorescence confocal microscopy was used to correlate distribution of Chit-1 and CHI3L1 expressing cells to TDP pathology.

**Results:** Chit-1 gene expression was increased in the spinal cord, and CHI3L1 expression was increased in both the spinal cord and motor cortex of sALS and C9-ALS patients when compared to controls. Highest levels of Chit-1^+^ glia were in cortical regions that contain hallmark neuropathology for each neurodegenerative disease. CHI3L1^+^ glia were only significantly increased in sALS. Neither Chit-1^+^ nor CHI3L1^+^ glia were in close proximity to pTDP containing neurons in the motor cortex gray matter; however, there was a significant co-localization of glial pTDP with Chit-1 and CHI3L1 in the motor cortex white matter.

**Conclusions:** Chit-1 and CHI3L1 expressing cells were most abundant in the white matter of cortical regions affected by each neurodegenerative disease and the spinal cord. Chit-1 or CHI3L1 expressing cells in the white matter also contained phosphorylated TDP-43. We also observed correlations between levels of Chit-1 or CHI3L1 expressing cells in the white matter to disease duration.

**KEY MESSAGES:** *What is already known on this topic:* Prior studies identified elevated levels of Chit-1 and CHI3L1 proteins in the CSF of various neurodegenerative conditions, though few studies examined levels of Chit-1 and CHI3L1 expressing cells both spatially and in relation to disease pathology.

*What this study adds:* We performed an extensive spatial characterization of Chit-1 and CHI3L1 protein levels across multiple regions and neurodegenerative conditions. This study also correlates Chit-1 and CHI3L1 expression to TDP pathology and other clinical parameters of disease duration.

*How this study might affect research, practice or policy:* Our findings indicate that the majority of Chit-1 and CHI3L1 expressing glia are located in the cortical subpial layer and the white matter, suggesting a role for chitinases in modulating neuroinflammatory mechanisms or reparative/regenerative responses in the white matter of ALS and other neurodegenerative diseases. This study suggests new therapeutic opportunities for targeting chitinase expressing cells in neurodegenerative diseases.

## BACKGROUND

Amyotrophic lateral sclerosis (ALS) is a progressive and fatal neurodegenerative disorder that affects both upper and lower motor neurons in the motor cortex, brainstem, and spinal cord [1–3]. There are several molecular mechanisms proposed to contribute to ALS, including neuroinflammation [4]. This neuroinflammatory process is predominantly mediated by glial cells in the central nervous system (CNS) that secrete many proteins, including cytokines, chemokines, and reactive oxygen species [5]. Activated glial cells release pro- and anti-inflammatory factors that impact local and distal cell types and ultimately can be measured in the cerebrospinal fluid (CSF) and blood [6]. In addition, recent studies have identified multiple glial subtypes with spatial and temporal heterogeneity during disease states, adding to the complexity of glial function and challenges in drug development [6–8]. One class of inflammatory modulators released by glial cells are chitinases, which have been shown to be elevated in the CSF of ALS and other neurodegenerative diseases [9, 10].

Human chitinases and chitinase-like proteins/chilectins, which belong to the glycosyl hydrolase family 18 proteins, are responsible for hydrolyzing chitin and chitin-containing pathogens, mediating the innate immune response, and aiding in tissue remodeling in mammals [11]. Chitinase-3-like protein 1 (CHI3L1 or YKL-40) and chitotriosidase (Chit-1) are two chitinases identified as biomarkers of ALS, with increased levels of both CHI3L1 and Chit-1 in the CSF of ALS patients when compared to healthy controls or other neurologic diseases [10, 12–17]. CSF levels of both CHI3L1 and Chit-1 also correlate with ALS disease progression, with significantly higher levels of CHI3L1 and Chit-1 in fast versus slow progressors [15]. Collectively, these studies implicate a role of chitinase proteins in ALS.

Cell types that express CHI3L1 or Chit-1 have been identified in postmortem tissues from both ALS and Alzheimer’s Disease (AD) patients. We detected CHI3L1 in subsets of astrocytes within the white matter of the motor cortex and spinal cord of ALS patients [15]. CHI3L1 immunoreactive astrocytes were also found in the frontal cortex of AD postmortem brain tissue, located near and surrounding amyloid beta plaques [18, 19]. Chit-1 expression has been reported in activated microglia in the spinal cord white matter of ALS patients [10]. However, no study has performed an extensive evaluation of CHI3L1 and Chit-1 expression in multiple brain regions and spinal cord across neurodegenerative diseases. We examined CHI3L1 and Chit-1 expression and cellular distribution in the motor cortex, frontal cortex, hippocampus, and lumbar spinal cord of ALS, AD, frontotemporal lobar dementia (FTLD), and non-neurologic disease controls (NNDCs). Our results indicate that CHI3L1 or Chit-1 expressing glia are most abundant in the white matter of affected brain and spinal cord regions across neurodegenerative diseases, supporting spatially specific regulated functions of chitinase expressing glia in these diseases.

## METHODS

### Subjects

Subject groups included those with clinical diagnosis and neuropathologic confirmation of sporadic ALS (sALS) lacking any known pathogenic gene variants, C9orf72 repeat expansion ALS (C9-ALS), FTLD, AD, or NNDCs. A total of 13 sALS, 12 C9-ALS, 12 FTLD, 14 AD, and 9 NNDC cases were used (Table 1). Postmortem human tissue samples were provided by the Target ALS Human Postmortem Tissue Core, NIH NeuroBioBank, and The Department of Veterans Affairs Biorepository Brain Bank. All participants provided IRB-approved informed consent for the collection of tissues and use in research studies.

**Table 1.**
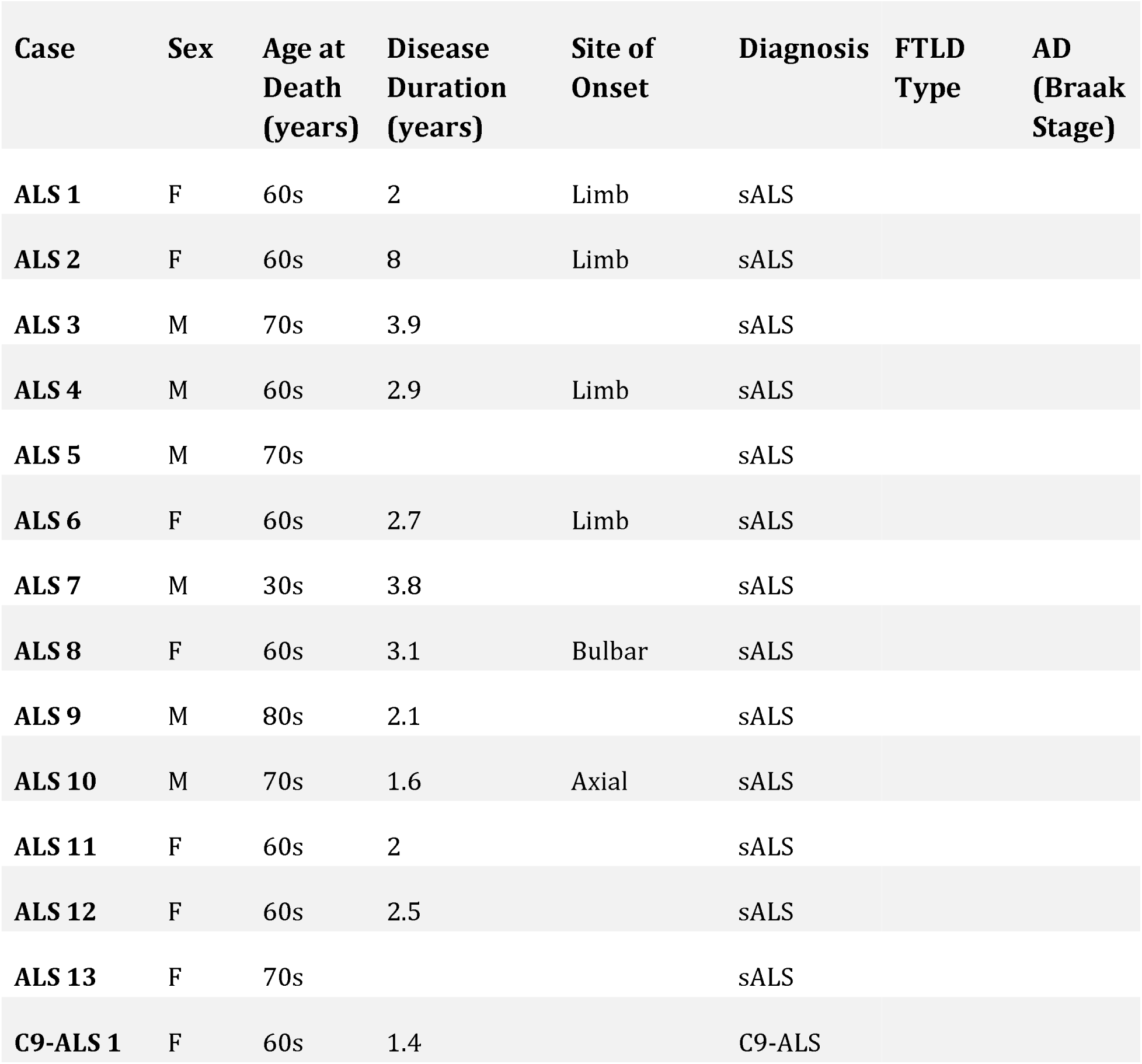
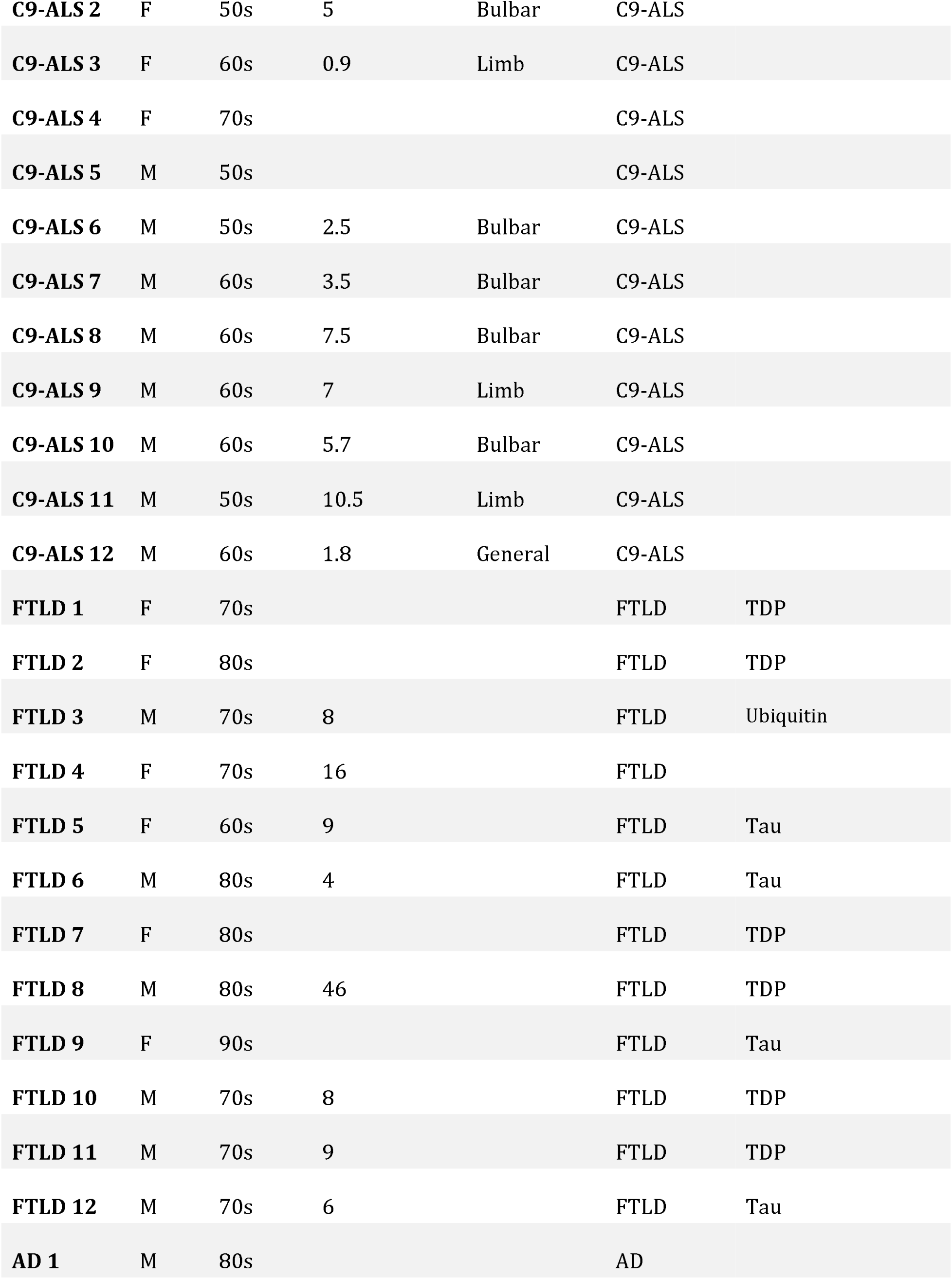
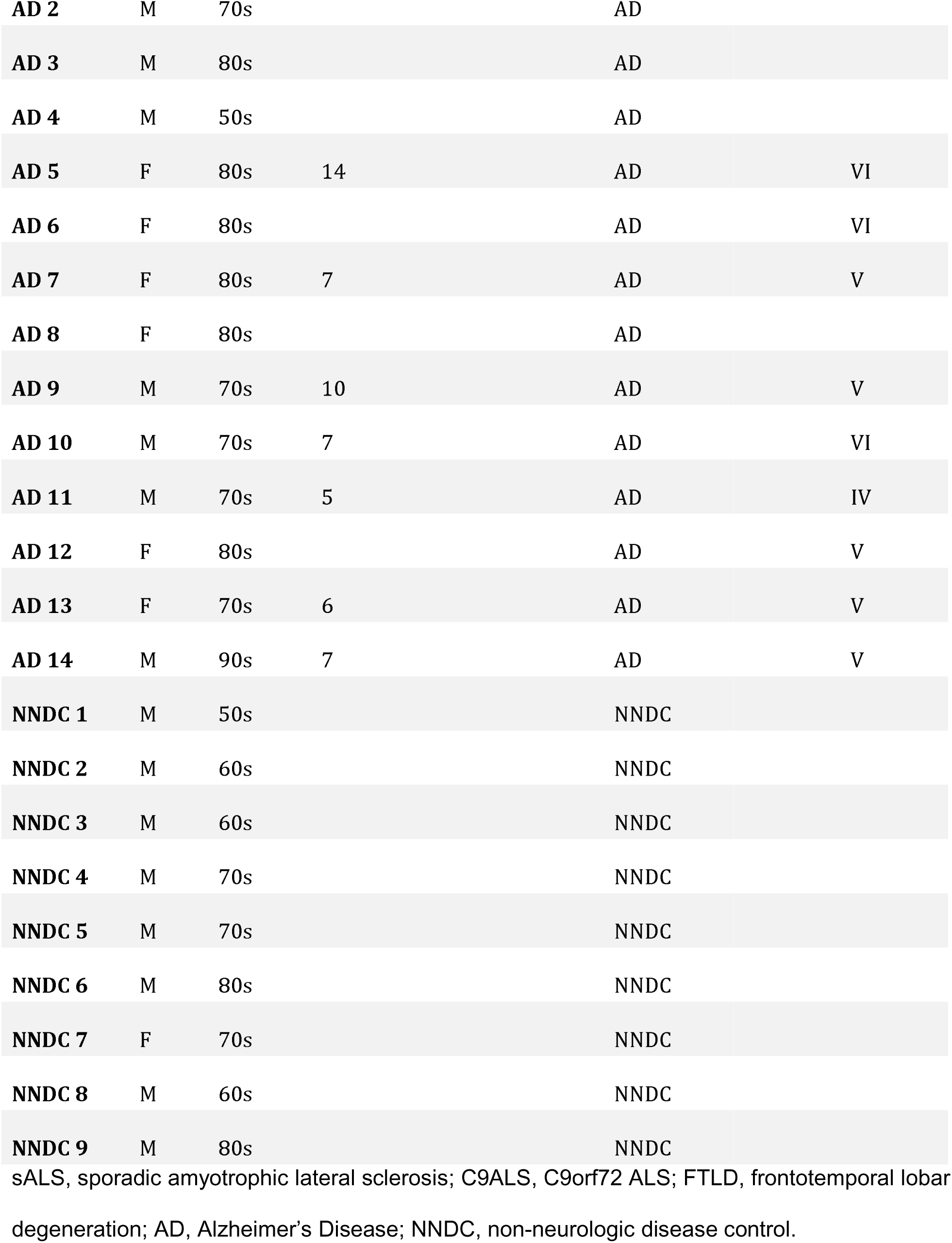
Subject Demographics.

### Bulk RNA-sequencing of Postmortem Tissue

Publicly available postmortem RNA sequencing data were obtained from the Target ALS Data Portal Postmortem Tissue Core (Target ALS, 2023) to identify transcriptomic levels of CHI3L1 and Chit-1 in sALS, C9-ALS, FTLD, AD, and NNDC participants. Transcriptomic profiles were obtained for the motor cortex [sALS (n=270), C9-ALS (n=33), and NNDC (n=46)] and lumbar spinal cord [sALS (n=140), C9-ALS (n=16), and NNDC (n=18)]. Counts were then normalized using size factor normalization. For visualization, dot plots were generated to perform comparative analyses of gene expression levels of CHI3L1 and CHIT1 across different conditions.

### Tissue Immunostaining

Immunohistochemistry (IHC) was performed on formalin-fixed paraffin-embedded (FFPE) tissue sections from motor cortex, frontal cortex, hippocampus, and lumbar spinal cord from sALS, C9-ALS, FTLD, AD, and NNDCs participants as previously described [15]. Slides were incubated with either rabbit anti-CHI3L1 (Thermo Fisher, Cat #: PA5-43746, 1:300) or goat anti-Chit-1 (R&D Systems, Cat #: AF3559, 2 µg/mL) primary antibodies. Sections were washed and subsequently incubated with the appropriate secondary antibody (biotinylated anti-rabbit or anti-goat IgG) (Vector Labs, 1:200) for one hour at room temperature. After washes, sections were incubated in the Vectastain Elite ABC HRP reagent (Vector Labs) for 30 minutes before being developed using the Vector ImmPACT NovaRED HRP substrate solution (Vector Labs) for two or three minutes. All sections were counterstained with Mayer’s haematoxylin (Sigma-Aldrich).

Images were acquired using the Leica Aperio VERSA Digital Pathology Scanner and analyzed using the Leica Aperio ImageScope software (version 12.3.2). Regions of interest (ROIs) were drawn for each slide encompassing layers 2-5 of the cortical gray matter, cortical white matter, CA1 and CA4 of the hippocampus, anterior horns and lateral corticospinal tracts of the spinal cord. ROIs were analyzed using the Aperio cytoplasmic algorithm where the following input parameters were defined: two visible stains (counterstain and biomarker). The algorithm was then trained to compute the color values for each corresponding stain using regions where the haematoxylin and the NovaRED were well-separated. Nuclei were identified using the following parameters: Type 0 (uses only the counterstain to detect nuclei) and exclusion of cells that were less than 10 µm^2^ in area. The upper limit threshold encompassed values from 0-255.

Cytoplasm segmentation was set based on the following parameters: 6µm maximum distance from the nuclei and three different thresholds for positive staining (1+, 2+, and 3+) (Figure S1). Positive thresholds encompassed values from 0-255 but were defined according to the level of background and architecture of each tissue. The percent positive cells and the percent of cells within each positivity threshold were calculated by the algorithm.

Tissue sections for immunofluorescent (IF) microscopy were deparaffinized, rehydrated, and epitope retrieval steps were performed as described above. Blocking and primary antibody incubation were performed in Superblock and then overnight at 4 , respectively. Slides were incubated in either rabbit anti-CHI3L1 (1:500) or goat anti-Chit-1 (1:500) and rat anti-phospho TDP43 (Biolegend, Cat #: SIG-39852, 1:500) primary antibodies. Following the primary antibody, slides were incubated in secondary antibodies donkey anti-Goat IgG Alexa Fluor 488, goat anti-rat IgG Alexa Fluor 555, goat anti-rabbit IgG Alexa Fluor 647, or goat anti-rabbit IgG Alexa Fluor 488 (Invitrogen, Carlsbad, California, USA, 1:200). All tissues were coverslipped using Vectashield Vibrance Antifade Mounting Medium with DAPI (Vector Labs).

Fluorescent images were acquired using a Nikon A1 HD25 confocal microscope. Five images per slide were taken for both the white and the gray matter of each tissue section. Images were taken at 60x magnification and analyzed using ImageJ to quantify co-localization of Chit-1 or CHI3L1 with phosphorylated TDP-43 (pTDP) aggregates and the amount of CHI3L1 immunoreactivity near Aβ plaques.

### Statistical Analyses

Statistical analysis was performed using GraphPad Prism V.10.2.3. All H scores calculated for IHC slides were normalized to the total cytoplasmic area prior to analysis. Kruskal-Wallis tests with a post hoc Dunn correction were performed to assess statistically significant differences of p<0.05 between each disease condition in the RNA-seq as well as the IHC datasets. Correlation of Chit-1 and CHI3L1 with age at death and disease duration was performed using Spearman’s rank correlation coefficient. Correlations were classified as follows unless otherwise noted: no correlation (0 - 0.39), moderate (0.4 - 0.59), strong (0.6 - 0.79), and very strong (0.8 - 1.0). Co-localization analysis of CHI3L1 and Chit-1 with glial pTDP was performed using Mander’s coefficients. The number of CHI3L1^+^ and Chit-1^+^ cells within 100µm of the neuronal pTDP aggregates was determined using the concentric circles plugin on ImageJ.

## RESULTS

### Chit-1 Expressing Cells in Affected Cortical and Spinal Cord Regions in Neurodegenerative Diseases

As noted above, Chit-1 expressing microglia were previously detected in the spinal cord posterior corticospinal tracts of ALS patients [10]. To confirm and expand upon this finding, we first examined Chit-1 gene expression profiles in the motor cortex and lumbar spinal cord of ALS, C9-ALS, and NNDC patients. We found significantly increased Chit-1 gene expression in the lumbar spinal cord of sALS and C9-ALS patients but not in the motor cortex (Figure 1A, B). We next examined Chit-1 protein expression in the motor cortex, frontal cortex, hippocampus, and spinal cord of ALS, C9-ALS, FTLD, AD, and NNDC patients by immunohistochemistry. We found that Chit-1 expressing cells (Chit-1^+^) were increased specifically in the motor cortex white matter of sALS relative to AD and NNDC cases (Figure 1C). Chit-1^+^ glia were also increased in the frontal cortex and hippocampal white matter of FTLD and the frontal cortex white matter of C9-ALS cases relative to sALS and NNDCs (Figure 1D, E). Additionally, we confirmed that Chit-1^+^ glia were elevated in sALS and C9-ALS spinal cord white matter and significantly increased when compared to NNDC cases (Figure 1F). Overall, there was a differential distribution of Chit-1^+^ glia in brain and spinal cord regions that contain hallmark neuropathology for each neurodegenerative disease.

**Figure 1:**
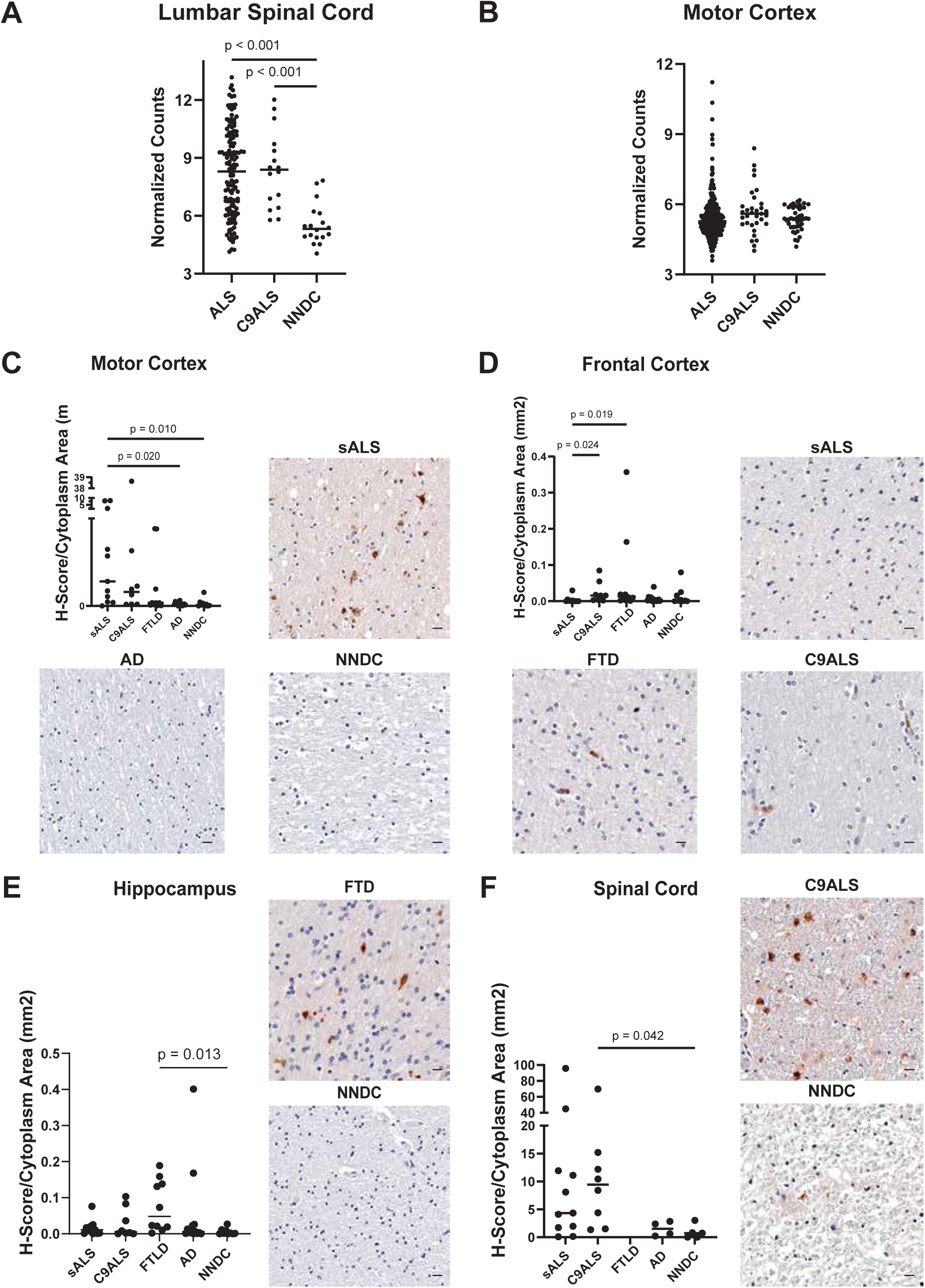
Chit-1 gene expression and immunohistochemistry in brain and spinal cord white matter. Normalized Chit-1 transcript counts from NYGC ALS Consortium bulk RNA-sequencing data in (A) lumbar spinal cord and (B) motor cortex. Pairwise comparisons by Kruskall-Wallis test, with *p* values corrected by Dunn post-hoc analysis. Lumbar spinal cord: sALS (n=140); C9ALS (n=16); NNDC (n=18). Motor cortex: sALS (n=270); C9ALS (n=33); NNDC (n=46). Representative images and quantification of Chit-1 positive cells in the white matter of the (C) motor cortex, (D) frontal cortex, (E) hippocampus, and (F) spinal cord. Data analysis was performed in a blinded manner as described in Methods. Pairwise comparisons by Kruskal-Wallis test with Dunn post-hoc correction were used to assess differences between subject groups for each cortical and spinal cord region. *P* values <0.05 shown. Chit-1, chitotriosidase-1; sALS, sporadic amyotrophic lateral sclerosis; C9ALS, C9orf72 ALS; FTLD, frontotemporal lobar degeneration; NNDC, non-neurologic disease control. All images at 20X magnification and scale bar = 10 µm.

We next quantified the intensity of Chit-1 immunostaining per cell to evaluate the relative level of protein expression within the white matter and compared these values across cortical and spinal cord regions. Cells that did not exhibit Chit-1 immunoreactivity were given a score of 0, and cells with Chit-1 protein expression were further defined as having weak (1+), moderate (2+), or strong (3+) immunoreactivity as described in the methods section. We found increased Chit-1^+^ glia in the motor cortex white matter of sALS and C9-ALS cases relative to AD, FTLD and NNDC, with C9-ALS having a higher percentage of moderate and strong immunoreactive cells than sALS (Figure 2A). FTLD cases exhibited the highest number of Chit-1+ cells in the white matter of the frontal cortex and hippocampus, though the total percent of white matter cells expressing Chit-1 was quite low (less than 1%) (Figure 2B, C). The lumbar spinal cord posterior corticospinal tracts in sALS and C9-ALS displayed the highest percentage of Chit-1^+^ glia, with sALS exhibiting higher staining intensity than C9-ALS (Figure 2D).

**Figure 2:**
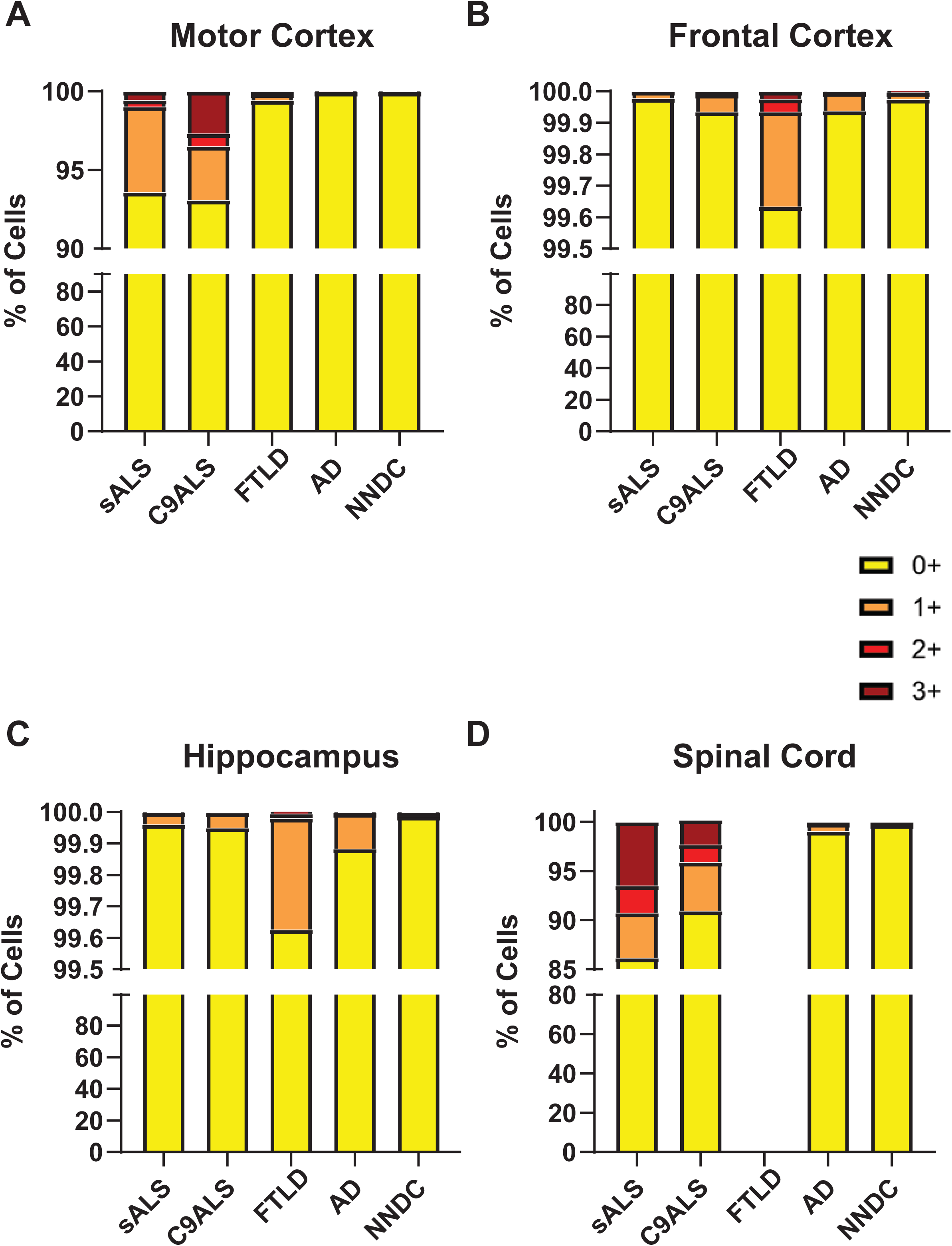
Percentage of cells expressing Chit-1 for each defined staining intensity (0+, 1+, 2+, 3+) in the white matter of the (A) motor cortex, (B) frontal cortex, (C) hippocampus, and (D) spinal cord of sALS, C9ALS, FTLD, AD, and NNDC patients. 0+: no expression (yellow); 1+: weak positive expression (orange); 2+: moderate positive expression (red); 3+: strong positive expression (burgundy). Chit-1, chitotriosidase-1; sALS, sporadic amyotrophic lateral sclerosis; C9ALS, C9orf72 ALS; FTLD, frontotemporal lobar degeneration; NNDC, non-neurologic disease control.

The mislocalization and aggregation of TDP-43 is a pathologic hallmark of sALS and most familial forms of ALS, as well as in significant numbers of AD and FTLD-TDP [20]. We assessed whether Chit-1^+^ microglia were in close proximity to neurons with TDP-43 pathology (pTDP) in sALS. Chit-1^+^ glia were not in close proximity to pTDP positive neurons within the motor cortex gray matter (Figure 3A). We also examined whether Chit-1 co-localizes with pTDP in microglia of the cortical white matter. We found that approximately 60% of Chit-1^+^ glia also contained pTDP (Figure 3B). Due to limitations in the number of C9-ALS and FTLD cases that have sufficient levels of both pTDP pathology and Chit-1 expressing microglia, we did not assess the localization of Chit-1 relative to pTDP pathology in these disease contexts. Overall, Chit-1^+^ microglia were not in close proximity to pTDP containing neurons in sALS motor cortex gray matter; however, there was a significant co-localization of glial pTDP and Chit-1 protein in the motor cortex white matter of sALS.

**Figure 3:**
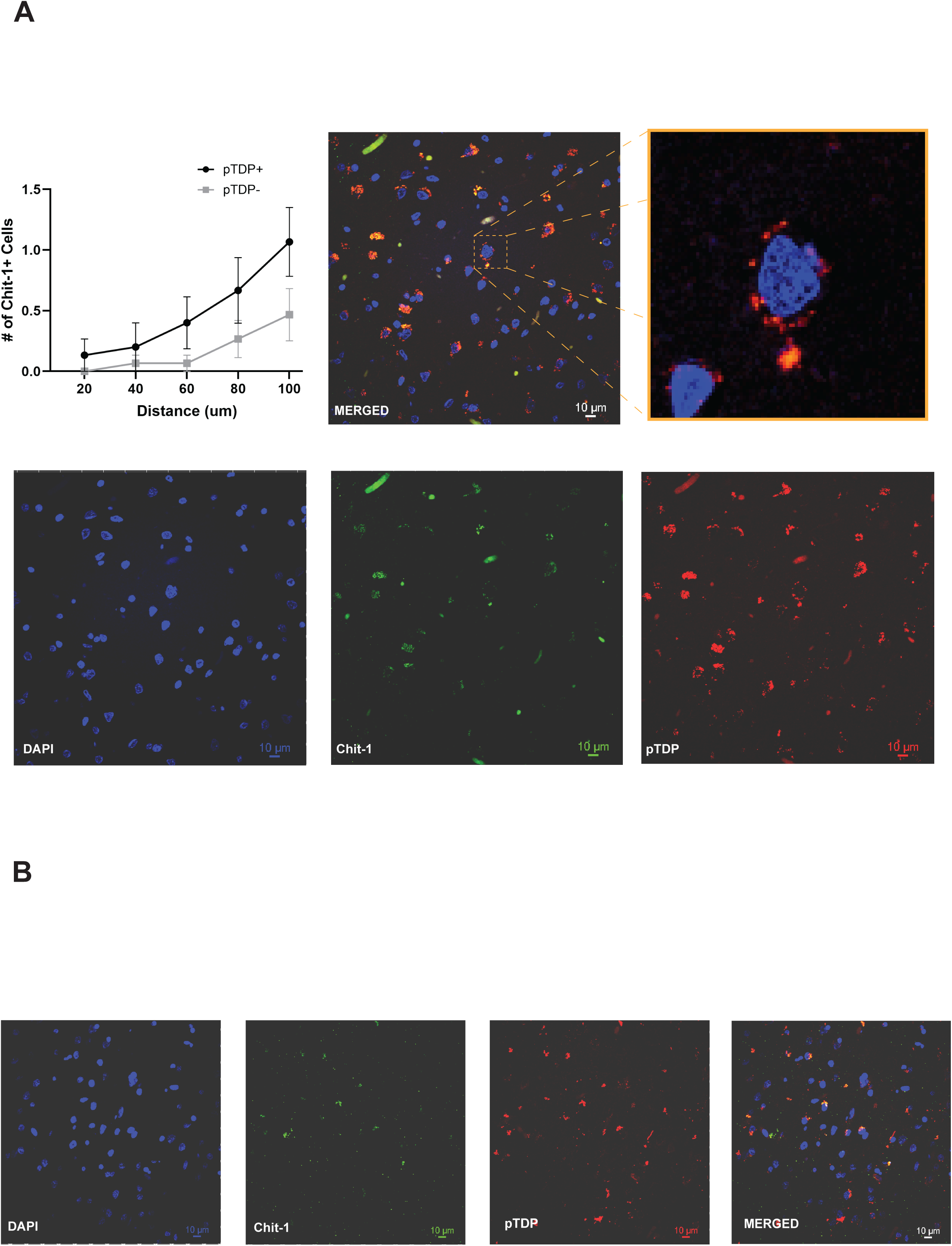
Immunofluorescence microscopy for Chit-1 and phosphorylated TDP-43 (pTDP) in sALS motor cortex. (A) Representative images used to measure distance between Chit-1 expressing cells and neuronal pTDP inclusions in the motor cortex gray matter using concentric curves. The zoomed image represents a neuron containing cytoplasmic pTDP and distance measured from center of neuron to Chit-1 expressing cells determined for 5 images per case. The graph represents the average distance of Chit-1 expressing cells to neurons containing (pTDP+) or lacking (pTDP-) pathology. (B) Co-localization of Chit-1 and pTDP positive glia in the motor cortex white matter. DAPI stained nuclei denoted in blue. Chit-1, chitotriosidase-1; sALS, sporadic amyotrophic lateral sclerosis. All images are 60x magnification and scale bar = 10 µm.

### CHI3L1 Expressing Cells in Affected Cortical and Spinal Cord Regions in Neurodegenerative Diseases

We next measured CHI3L1 gene expression in the motor cortex and lumbar spinal cord using bulk RNA-seq of postmortem tissue samples. CHI3L1 gene expression was significantly increased in both the lumbar spinal cord and motor cortex of sALS and C9-ALS patients relative to the NNDCs (Figure 4A, B).

**Figure 4:**
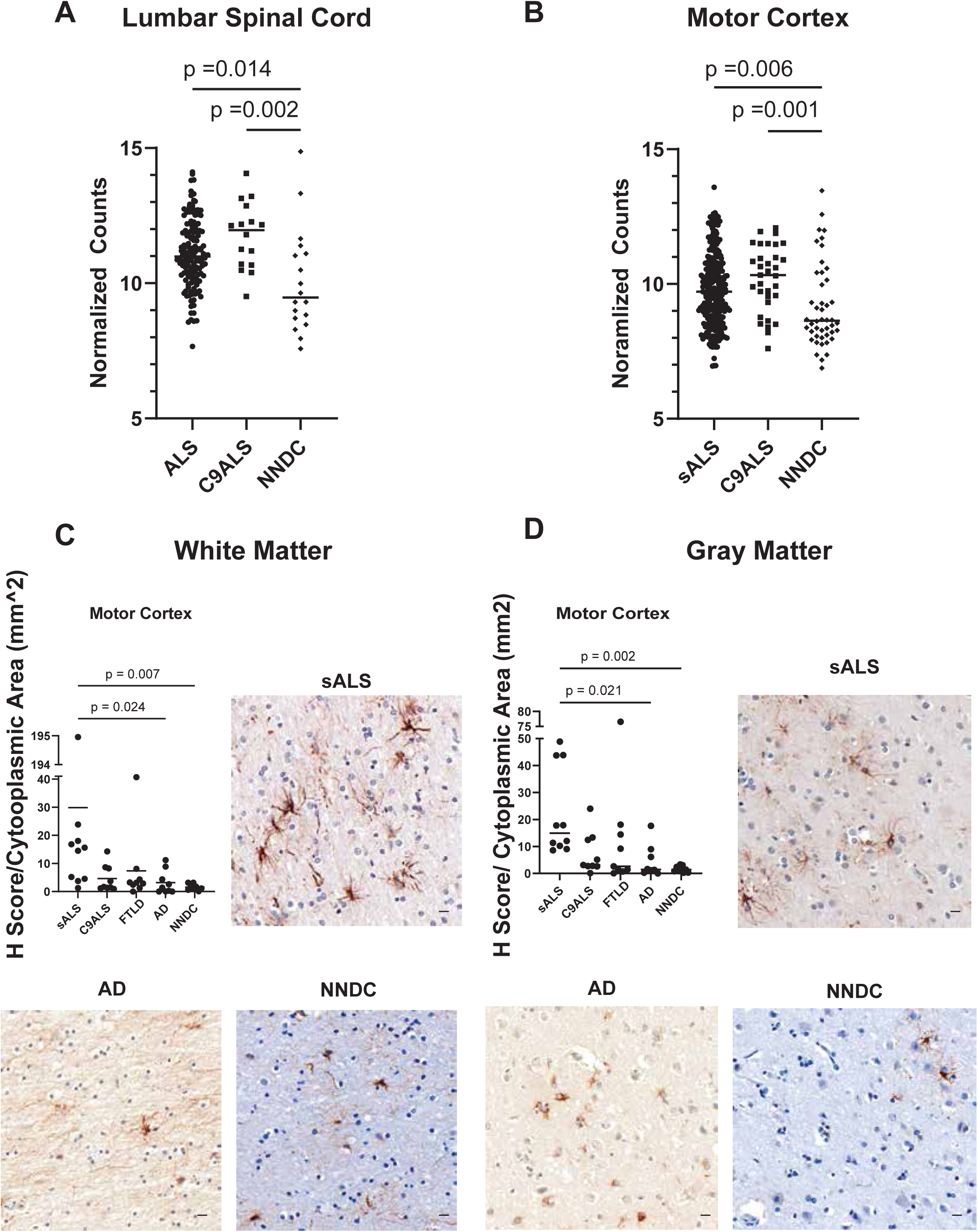
CHI3L1 gene expression and immunohistochemistry in brain and spinal cord white matter. Normalized CHI3L1 transcript counts from NYGC ALS Consortium bulk RNA-sequencing data in (A) lumbar spinal cord and (B) motor cortex. Pairwise comparisons by Kruskall-Wallis test, with *p* values corrected by Dunn post-hoc analysis. Lumbar spinal cord: sALS (n=140); C9ALS (n=16); NNDC (n=18). Motor cortex: sALS (n=270); C9ALS (n=33); NNDC (n=46). Representative images and quantification of CHI3L1 positive cells in the (C) white matter and (D) gray matter of the of the motor cortex. Data analysis was performed in a blinded manner as described in Methods. Pairwise comparisons by Kruskal-Wallis test with Dunn post-hoc correction were used to assess differences between subject groups for each cortical and spinal cord region. *P* values <0.05 shown. CHI3L1, Chitinase-3-like 1; sALS, sporadic amyotrophic lateral sclerosis; C9ALS, C9orf72 ALS; FTLD, frontotemporal lobar degeneration; NNDC, non-neurologic disease control. All images at 20X magnification and scale bar = 10 µm.

We previously reported CHI3L1 protein expression in a subset of activated astrocytes in the white matter and subpial layer of the motor cortex in sALS [15]. Here, we confirmed CHI3L1 immunoreactive cells (CHI3L1^+^) at the subpial layer and in the white matter of cortical brain regions in sALS, with increased CHI3L1^+^ glia in the motor cortex when compared to AD and NNDC cases (Figure 4C). Interestingly, there were also increased CHI3L1^+^ cells in sALS gray matter when compared to AD and NNDCs (Figure 4D). While there is a trend of higher numbers of CHI3L1+ cells in sALS versus C9-ALS, this did not reach statistical significance. One FTLD case exhibited increased CHI3L1^+^ cells in the motor cortex white and gray matter, but overall values across all FTLD cases were not increased when compared to AD or NNDC. CHI3L1^+^ cells were not significantly altered across neurodegenerative diseases in the frontal cortex, hippocampus, or spinal cord (Figure S4). Overall, CHI3L1 showed differential immunoreactivity only in astrocytes of the motor cortex.

CHI3L1 protein levels were also qualitatively assessed by immunoreactivity within each cell. Cells that did not express CHI3L1 were given a score of 0, and CHI3L1 immunoreactive cells were scored as follows: weak (1+), moderate (2+), or strong (3+) immunoreactivity. The percentage of CHI3L1^+^ cells in the white matter was much higher overall across all CNS regions and disease conditions compared to the percentage of white matter cells expressing Chit-1 (Figure 5). CHI3L1^+^ cells were more prevalent in the motor and frontal cortex in diseases that fall along the ALS-FTD spectrum. CHI3L1 immunoreactive cells that exhibited strong expression were most prevalent in the motor cortex of sALS and the frontal cortex of FTLD cases (Figure 5A, B). In the hippocampus, CHI3L1 positive cells were evident in the white matter of all cases, but the cells with strong positive expression were most prevalent in C9-ALS and FTLD cases (Figure 5C). Surprisingly, CHI3L1 positive cells in the spinal cord were most abundant in the small number of AD cases (Figure 5D), though this finding must be confirmed with larger numbers of AD spinal cord samples.

**Figure 5:**
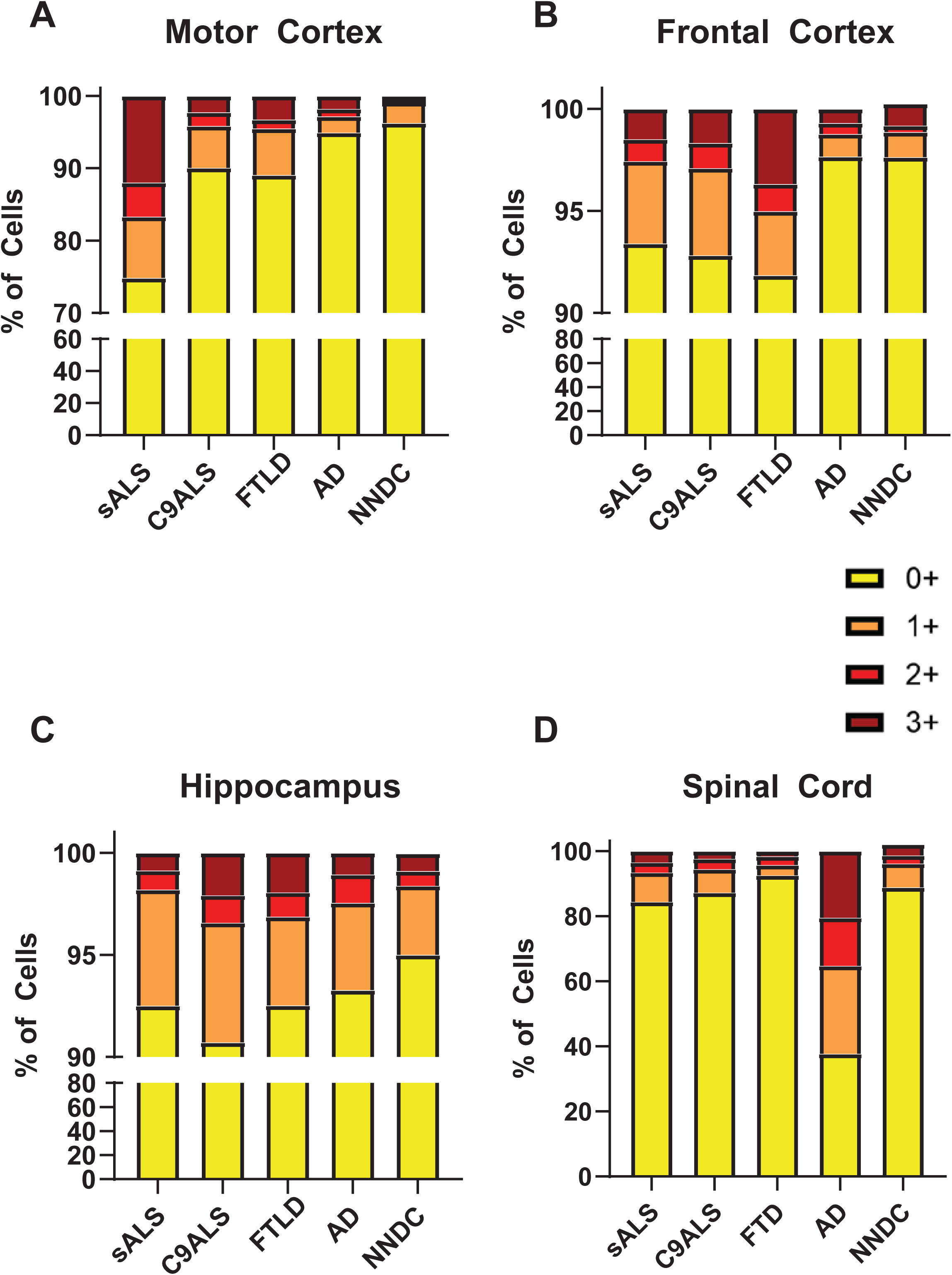
Percentage of cells expressing CHI3L1 for each defined staining intensity (0+, 1+, 2+, 3+) in the white matter of the (A) motor cortex, (B) frontal cortex, (C) hippocampus, and (D) spinal cord of sALS, C9ALS, FTLD, AD, and NNDC patients. 0+: no expression (yellow), 1+: weak positive expression (orange), 2+: moderate positive expression (red), 3+: strong positive expression (burgundy). CHI3L1, Chitinase-3-like 1; sALS, sporadic amyotrophic lateral sclerosis; C9ALS, C9orf72 ALS; FTD, frontotemporal lobar degeneration; NNDC, non-neurologic disease control.

We next assessed the spatial distribution of CHI3L1 immunoreactivity to neurons with pTDP pathology in the motor cortex gray matter in sALS and C9-ALS and the frontal cortex in FTLD. We did not detect CHI3L1^+^ astrocytes to be discriminately localized near pTDP containing neurons in sALS, C9-ALS, or FTD in the gray matter (Figure 6A, B, C). In the white matter, approximately 60% of CHI3L1^+^ astrocytes contained pTDP (Figure 6D).

**Figure 6:**
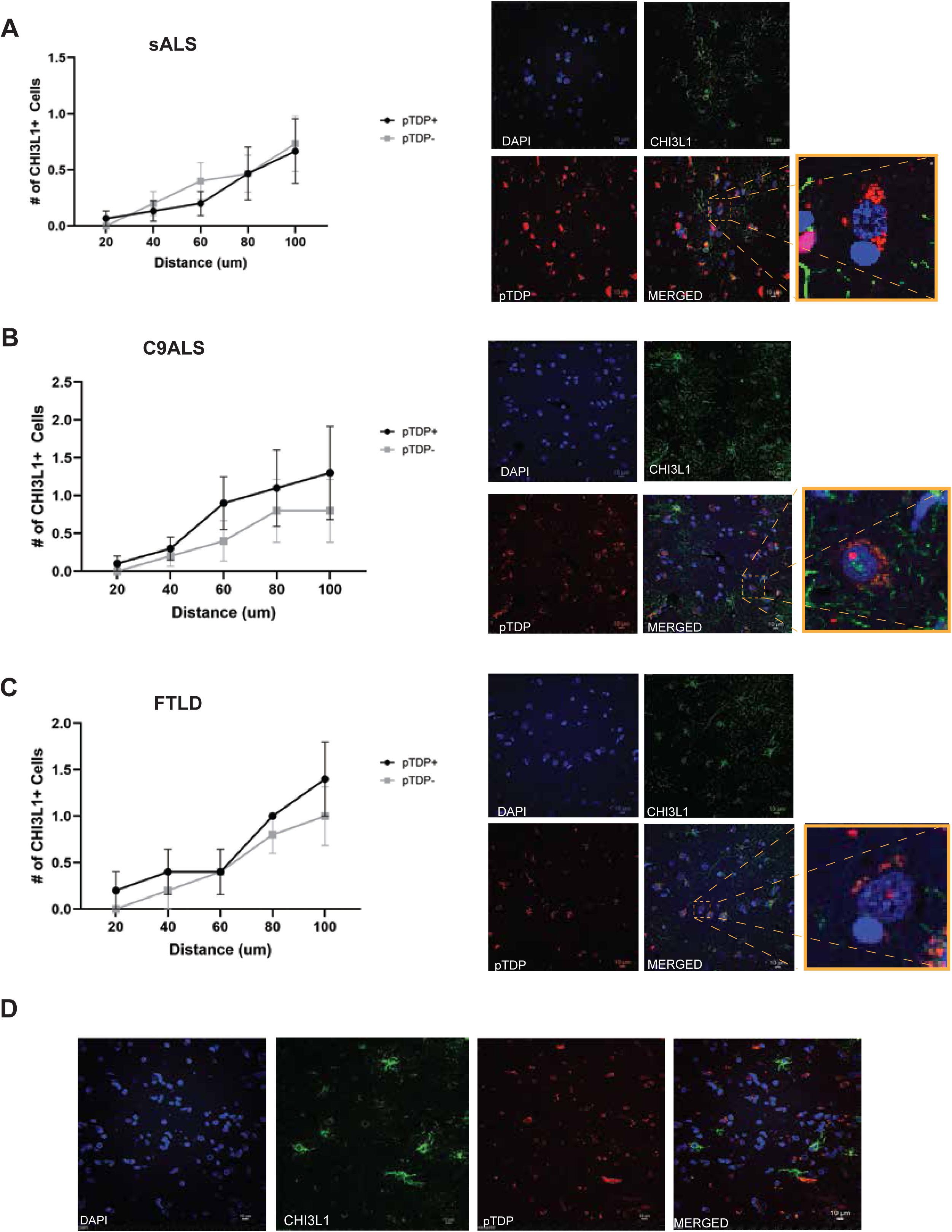
Immunofluorescence microscopy for CHI3L1 and phosphorylated TDP-43 (pTDP) in gray matter of (A) sALS motor cortex, (B) C9-ALS motor cortex, and (C) frontal cortex of FTLD. relative to neurons with (pTDP+) or without (pTDP-) pathological TDP inclusions. Representative images are shown to measure distance between CHI3L1 expressing cells and neuronal pTDP inclusions using concentric curves. The zoomed image represents a neuron containing cytoplasmic pTDP and distance measured from center of neuron to CHI3L1 expressing cells determined for 5 images per case. Each graph represents the average distance of CHI3L1 expressing cells to neurons containing (pTDP+) or lacking (pTDP-) pathology. (D) Co-localization of CHI3L1 and pTDP positive glia in the motor cortex white matter. DAPI stained nuclei denoted in blue. CHI3L1, Chitinase-3-like 1; sALS, sporadic amyotrophic lateral sclerosis; C9ALS, C9orf72 ALS; FTLD, frontotemporal lobar degeneration. All images are 60x magnification and scale bar = 10 µm.

### Correlation of Chit-1+ and CHI3L1+ Cells to Clinical Parameters of Disease

Prior studies reported that CSF levels of Chit-1 and CHI3L1 protein correlate with the rate of ALS disease progression [15, 21–23]. Here we correlated levels of Chit-1 and CHI3L1 expressing cells in the white and gray matter of the motor cortex and spinal cord with each other, the age of disease onset, and disease duration for sALS, C9-ALS, and FTLD cases. We found that for sALS, there was a moderate negative correlation between Chit-1^+^ cells in the gray matter of the spinal cord and the white matter of the motor cortex with disease duration (Figure 7A). There was a moderate positive correlation of Chit-1+ cells in the spinal cord white matter to age at death, as well as motor cortex gray matter to both motor cortex and spinal cord white matter (Figure 7A). In C9-ALS, there was a moderate negative correlation between Chit-1^+^ cells in the white matter of the spinal cord and disease duration, while motor cortex white matter Chit-1^+^ cells showed a moderate negative correlation to age at death (Figure 7B). Levels of Chit-1^+^ cells in the gray and white matter of the spinal cord were strongly correlated with each other, and Chit-1^+^ cells in the gray and white matter of the motor cortex were moderately correlated in both sALS and C9-ALS (Figure 7A, B). The levels of Chit-1+ cells were strongly correlated between the white and gray matter in the frontal cortex of FTLD cases but were not correlated to disease duration or age at death (Figure 7C).

**Figure 7:**
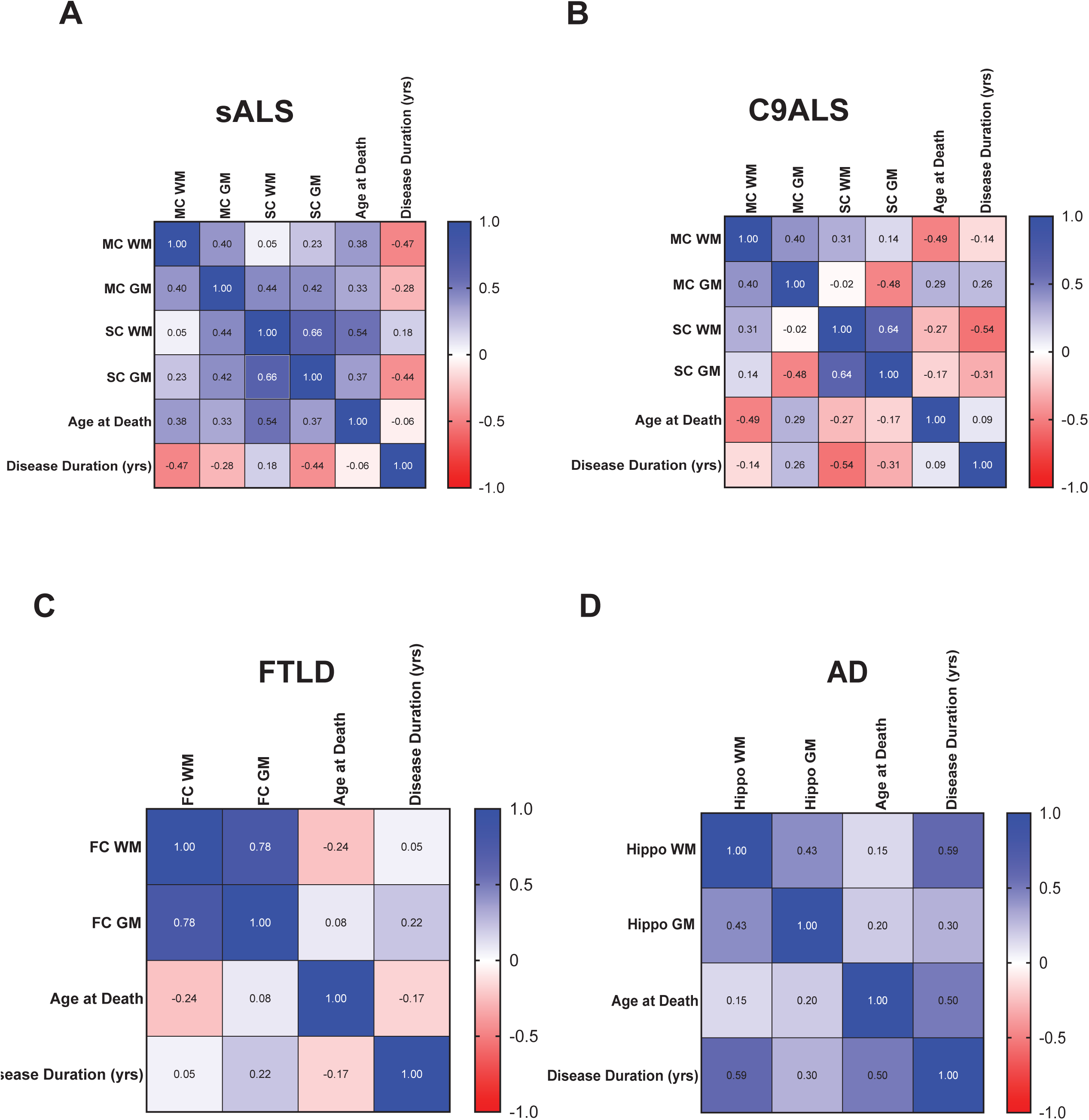
Correlation matrix of Chit-1-positive cells in the white and gray matter of the motor cortex, lumbar spinal cord, age at death, and disease duration in (A) sALS and (B) C9-ALS. (C) Correlation matrix of Chit-1-positive cells in the white and gray matter of the frontal cortex, age at death, and disease duration in FTLD. (D) Correlation matrix of Chit-1-positive cells in the white and gray matter of the hippocampus, age at death and disease duration in AD. Chit-1, chitotriosidase-1; sALS, sporadic amyotrophic lateral sclerosis; C9ALS, C9orf72 ALS; FTLD, frontotemporal lobar degeneration; AD, Alzheimer’s Disease.

Within AD hippocampus, Chit-1+ cells within the white matter were moderately correlated to disease duration, but levels between the white and gray matter did not strongly correlate (Figure 7D).

Interestingly, CHI3L1^+^ cells in the gray and white matter of the motor cortex showed a strong negative correlation with disease duration in sALS but a strong positive correlation to disease duration in C9-ALS (Figure 8A,B). Levels of CHI3L1^+^ cells were strongly correlated in the gray and white matter in the spinal cord but not in the motor cortex in sALS, while no correlations were observed between the white and gray matter in C9-ALS (Figure 8A,B). We also observed a moderate positive correlation between spinal cord gray matter and disease duration in sALS and a strong negative correlation between motor cortex and spinal cord white matter (Figure 8A,B). CHI3L1^+^ cells in FTLD frontal cortex gray matter exhibited a strong correlation to disease duration, but there was no correlation between levels in the white and gray matter (Figure 8C). CHI3L1^+^ cells in the hippocampus white and gray matter strongly correlated to each other, while the gray matter levels were negatively correlated to disease duration (Figure 8D).

**Figure 8:**
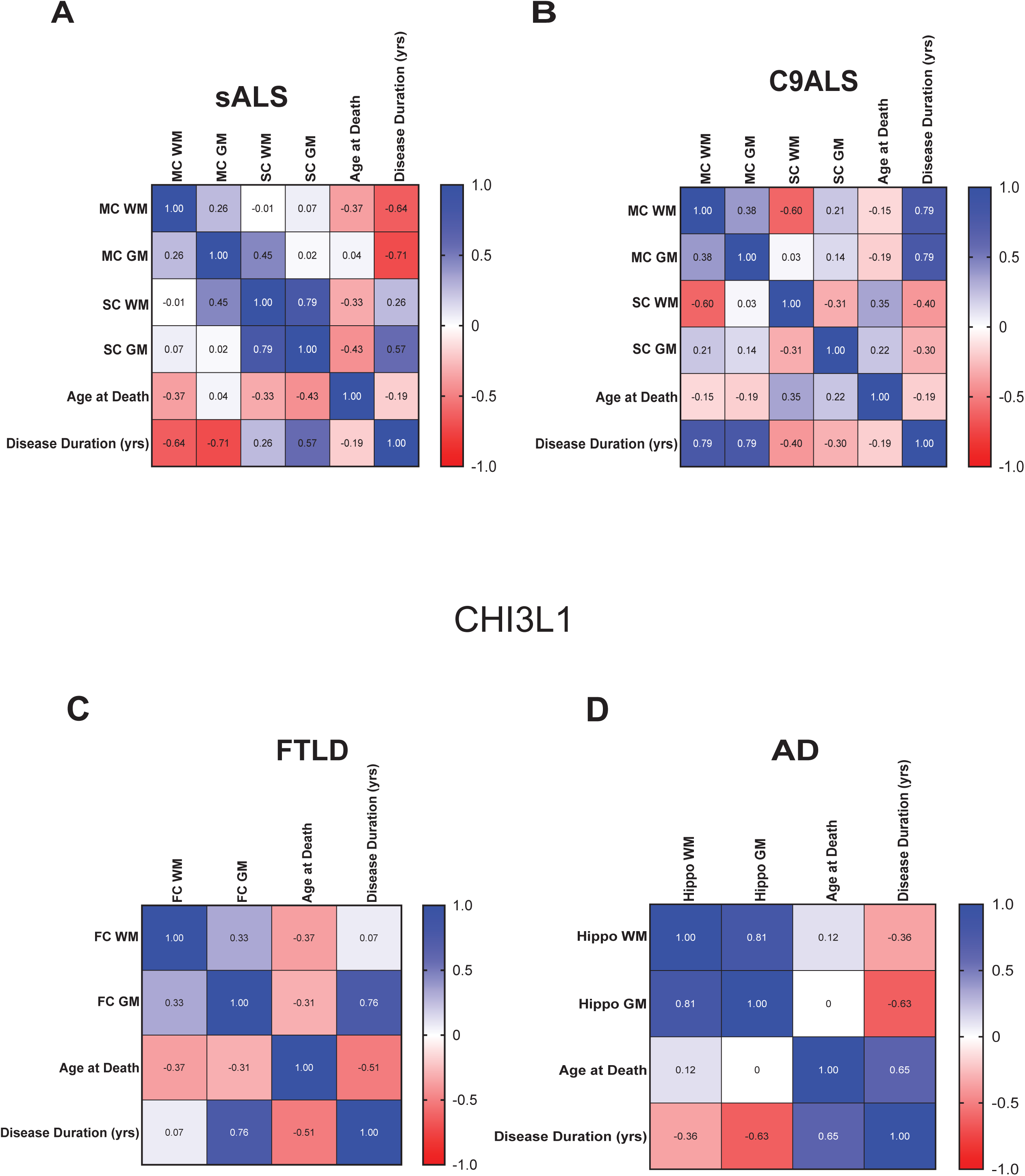
Correlation matrix of CHI3L1-positive cells in the white and gray matter of the motor cortex, lumbar spinal cord, age at death, and disease duration in (A) sALS and (B) C9-ALS. (C) Correlation matrix of CHI3L1-positive cells in the white and gray matter of the frontal cortex, age at death, and disease duration in FTLD. (D) Correlation matrix of CHI3L1-positive cells in the white and gray matter of the hippocampus, age at death and disease duration in AD. CHI3L1, Chitinase-3-like 1; sALS, sporadic amyotrophic lateral sclerosis; C9ALS, C9orf72 ALS; FTLD, frontotemporal lobar degeneration; AD, Alzheimer’s Disease.

## DISCUSSION

Prior studies have reported increased levels of Chit-1 and CHI3L1 (YKL-40) in the CSF of ALS and other neurodegenerative diseases and specific glial subtypes that express either Chit-1 or CHI3L1 [9, 10, 12–17]. In this study, we further characterized the abundance and distribution of cells expressing either Chit-1 or CHI3L1 in postmortem tissue from the ALS-FTD spectrum and AD. We observed that Chit-1+ and CHI3L1+ glia were differentially distributed across neurodegenerative diseases, with focal localization to white matter (subpial surface and white matter tracks) in brain and spinal cord regions that contain hallmark neuropathology associated with ALS, C9-ALS, FTLD, or AD.

We also explored the spatial distribution of chitinase expressing glia to neuronal TDP-43 pathology in gray matter and co-localization of chitinase protein and pTDP-43 in white matter. The percentage of cells expressing Chit-1 in the gray matter across neurodegenerative diseases was low (Figure S3), so we focused efforts on the motor cortex of sALS and C9-ALS cases which exhibit more consistent and evident pTDP-43 pathology. Within the gray matter, there was no spatial correlation between Chit-1+ cells and neurons containing pTDP-43. However, we did find that over 50% of Chit-1+ glia in the white matter of the motor cortex also contain pTDP-43 (Figure 3). Further studies are required to determine the temporal pattern of this co-localization and if altered TDP-43 function contributes to Chit-1 expression in glia. The overall percentage of CHI3L1+ cells in the gray matter was higher than Chit-1 across the various regions and neurodegenerative diseases (Figure S4 and S5). Previously, it was shown that CHI3L1+ astrocytes were located near amyloid beta plaques in the hippocampus of AD [18, 19].

However, we failed to detect a spatial correlation between CHI3L1+ cells and neurons containing pTDP-43 in the motor cortex gray matter of sALS or C9-ALS patients. We observed co-localization of pTDP-43 in 60-80% of CH3L1+ cells in the motor cortex white matter of sALS and C9-ALS patients. These findings again suggest a link between TDP-43 phosphorylation and chitinase expression in glial cells.

We also correlated levels of chitinase expressing cells in the white and gray matter with disease duration and age of death for sALS, C9-ALS, FTLD and AD. For Chit-1+ cells, we observed a weak negative correlation between number of Chit-1+ cells in the motor cortex white matter and disease duration but a modest negative correlation between Chit-1+ cells in the spinal cord white matter and age at death in sALS (Figure 7). In C9-ALS Chit-1+ cells exhibited a negative correlation to disease duration in the spinal cord white matter. We found no correlation between Chit-1+ cells in the frontal cortex and disease duration or age of death in FTLD but did observe a moderate positive correlation of Chit-1+ cells in the hippocampal white matter with disease duration (Figure 7D). These results suggest that higher levels of Chit-1+ cells in the white matter of the motor cortex of sALS or C9-ALS correlates to reduced disease duration or age of death, like that reported for CSF levels of Chit-1 in ALS patients. In AD, we detected a positive correlation of Chit-1+ cells in the hippocampal white matter to disease duration, suggesting a more protective effect of these cells to increase disease duration. Across all neurodegenerative diseases, we often observed a positive correlation between the levels of Chit-1+ cells in the white and gray matter in the regions predominately impacted by neuropathology.

For correlations between CHI3L1+ cells and disease duration, we found interesting and opposite results in sALS and C9-ALS. Levels of CHI3L1+ cells in the motor cortex white and gray matter were negatively correlated to disease duration in sALS but positively correlated to disease duration in C9-ALS (Figure 8). These results indicate that a higher number of CHI3L1+ cells in the motor cortex of C9-ALS patients increase disease duration (slow disease progression) but have the opposite impact in sALS patients (reduce disease duration). It has been reported that CHI3L1 can stimulate the proliferation of oligodendrocyte precursor cells as a method of myelin formation [24, 25], so it is possible that elevated levels of CHI3L1+ cells are a compensatory mechanism for repairing damaged axons in C9-ALS. However, these cells fail to induce the same compensatory impact in sALS. The underlying mechanisms contributing to this observation may be related to specific subtypes of glia expressing CHI3L1 in this familial form of ALS and are worthy of further investigation. Alternatively, disease progression and duration in C9-ALS may be driven more by motor cortex mechanisms and pathobiology than sALS. Similarly, we observed a positive correlation between CHI3L1+ cells in the frontal cortex gray matter of FTLD patients and disease duration, again suggesting a beneficial impact that impedes disease progression.

One limitation of our study is that multiple cell types can express each of the chitinase proteins. Both Chit-1 and CHI3L1 may be expressed in microglia, astrocytes, neutrophils, macrophages, and oligodendrocytes under the proper conditions. Single cell RNA-seq or spatial transcriptomics within tissue sections across these neurodegenerative diseases will provide important insights into the subtype of glial cells expressing each chitinase protein in the gray and white matter.

Overall, our study characterizes the expression and distribution of Chit-1+ and CHI3L1+ cells in brain and spinal cord regions of ALS, FTLD and AD, impacted by neuropathologic hallmarks of each neurodegenerative disease. Both Chit-1 and CHI3L1 expressing cells are most dominant in the subpial layer and white matter tracts in each neurodegenerative disease. We correlated levels of chitinase expressing cells to disease duration and found that CHI3L1+ cells in the motor cortex were positively correlated with disease duration for C9-ALS but negatively correlated with disease duration in sALS. These results suggest different pathogenic mechanisms within the motor cortex contributing to overall disease course. Continued studies are necessary to further define the mechanistic role of each chitinase protein and exact subtypes of glia within the white and gray matter that express each chitinase protein in neurodegenerative diseases.

## Data Availability

All data produced in the present work are contained in the manuscript.

## Acknowledgements

The authors thank all participants and their families for consenting to provide postmortem tissues used in this study. Tissue biobanks included the Target ALS Postmortem Tissue Core, NIH NeuroBioBank, and the Department of Veterans Affairs Biorepository Brain Bank. We thank the NYGC ALS Consortium for providing the bulk tissue transcriptomic data used in this study. Funding support was provided by the Barrow Neurological Foundation and Target ALS grant CF-2025-PMT to RB.

## Notes

### Competing Interest Statement

RB reports receiving consulting fees from Amylyx, Alector LLC, AcuraStem, Clene Therapeutics, NeuroSense Therapeutics, and BrainStorm, and is a co-founder and has stock options in nVector, Inc.

### Author Declarations

IRB of Barrow Neurological Institute gave ethical approval for this work (IRB Protocol #: PHXB-13BN080).

## References

1. Kiernan MC, Vucic S, Cheah BC, Turner MR, Eisen A, Hardiman O, et al. Amyotrophic lateral sclerosis. Lancet. 2011;377(9769):942–55.

2. Balendra R, Jones A, Jivraj N, Knights C, Ellis CM, Burman R, et al. Estimating clinical stage of amyotrophic lateral sclerosis from the ALS Functional Rating Scale. Amyotroph Lateral Scler Frontotemporal Degener. 2014;15(3-4):279–84.

3. Hardiman O, Al-Chalabi A, Chio A, Corr EM, Logroscino G, Robberecht W, et al. Amyotrophic lateral sclerosis. Nat Rev Dis Primers. 2017;3:17085.

4. Kwon HS, Koh SH. Neuroinflammation in neurodegenerative disorders: the roles of microglia and astrocytes. Transl Neurodegener. 2020;9(1):42.

5. DiSabato DJ, Quan N, Godbout JP. Neuroinflammation: the devil is in the details. J Neurochem. 2016;139 Suppl 2(Suppl 2):136–53.

6. Thonhoff JR, Simpson EP, Appel SH. Neuroinflammatory mechanisms in amyotrophic lateral sclerosis pathogenesis. Curr Opin Neurol. 2018;31(5):635–9.

7. Martins-Ferreira R, Calafell-Segura J, Leal B, Rodriguez-Ubreva J, Martinez-Saez E, Mereu E, et al. The Human Microglia Atlas (HuMicA) unravels changes in disease-associated microglia subsets across neurodegenerative conditions. Nat Commun. 2025;16(1):739.

8. Miller SJ, Philips T, Kim N, Dastgheyb R, Chen Z, Hsieh YC, et al. Molecularly defined cortical astroglia subpopulation modulates neurons via secretion of Norrin. Nat Neurosci. 2019;22(5):741–52.

9. Hall S, Surova Y, Ohrfelt A, Swedish Bio FS, Blennow K, Zetterberg H, Hansson O. Longitudinal Measurements of Cerebrospinal Fluid Biomarkers in Parkinson’s Disease. Mov Disord. 2016;31(6):898–905.

10. Steinacker P, Verde F, Fang L, Feneberg E, Oeckl P, Roeber S, et al. Chitotriosidase (CHIT1) is increased in microglia and macrophages in spinal cord of amyotrophic lateral sclerosis and cerebrospinal fluid levels correlate with disease severity and progression. J Neurol Neurosurg Psychiatry. 2018;89(3):239–47.

11. Pinteac R, Montalban X, Comabella M. Chitinases and chitinase-like proteins as biomarkers in neurologic disorders. Neurol Neuroimmunol Neuroinflamm. 2021;8(1).

12. Varghese AM, Sharma A, Mishra P, Vijayalakshmi K, Harsha HC, Sathyaprabha TN, et al. Chitotriosidase - a putative biomarker for sporadic amyotrophic lateral sclerosis. Clin Proteomics. 2013;10(1):19.

13. Collins MA, An J, Hood BL, Conrads TP, Bowser RP. Label-Free LC-MS/MS Proteomic Analysis of Cerebrospinal Fluid Identifies Protein/Pathway Alterations and Candidate Biomarkers for Amyotrophic Lateral Sclerosis. J Proteome Res. 2015;14(11):4486–501.

14. Thompson AG, Gray E, Bampton A, Raciborska D, Talbot K, Turner MR. CSF chitinase proteins in amyotrophic lateral sclerosis. J Neurol Neurosurg Psychiatry. 2019;90(11):1215–20.

15. Vu L, An J, Kovalik T, Gendron T, Petrucelli L, Bowser R. Cross-sectional and longitudinal measures of chitinase proteins in amyotrophic lateral sclerosis and expression of CHI3L1 in activated astrocytes. J Neurol Neurosurg Psychiatry. 2020;91(4):350–8.

16. Gray E, Thompson AG, Wuu J, Pelt J, Talbot K, Benatar M, Turner MR. CSF chitinases before and after symptom onset in amyotrophic lateral sclerosis. Ann Clin Transl Neurol. 2020;7(8):1296–306.

17. Gaur N, Perner C, Witte OW, Grosskreutz J. The Chitinases as Biomarkers for Amyotrophic Lateral Sclerosis: Signals From the CNS and Beyond. Front Neurol. 2020;11:377.

18. Craig-Schapiro R, Perrin RJ, Roe CM, Xiong C, Carter D, Cairns NJ, et al. YKL-40: a novel prognostic fluid biomarker for preclinical Alzheimer’s disease. Biol Psychiatry. 2010;68(10):903–12.

19. Moreno-Rodriguez M, Perez SE, Nadeem M, Malek-Ahmadi M, Mufson EJ. Frontal cortex chitinase and pentraxin neuroinflammatory alterations during the progression of Alzheimer’s disease. J Neuroinflammation. 2020;17(1):58.

20. Jo M, Lee S, Jeon YM, Kim S, Kwon Y, Kim HJ. The role of TDP-43 propagation in neurodegenerative diseases: integrating insights from clinical and experimental studies. Exp Mol Med. 2020;52(10):1652–62.

21. Thompson AG, Gray E, Thezenas ML, Charles PD, Evetts S, Hu MT, et al. Cerebrospinal fluid macrophage biomarkers in amyotrophic lateral sclerosis. Ann Neurol. 2018;83(2):258–68.

22. Andres-Benito P, Dominguez R, Colomina MJ, Llorens F, Povedano M, Ferrer I. YKL40 in sporadic amyotrophic lateral sclerosis: cerebrospinal fluid levels as a prognosis marker of disease progression. Aging (Albany NY). 2018;10(9):2367–82.

23. Gille B, De Schaepdryver M, Dedeene L, Goossens J, Claeys KG, Van Den Bosch L, et al. Inflammatory markers in cerebrospinal fluid: independent prognostic biomarkers in amyotrophic lateral sclerosis? J Neurol Neurosurg Psychiatry. 2019;90(12):1338–46.

24. Starossom SC, Campo Garcia J, Woelfle T, Romero-Suarez S, Olah M, Watanabe F, et al. Chi3l3 induces oligodendrogenesis in an experimental model of autoimmune neuroinflammation. Nat Commun. 2019;10(1):217.

25. Jiang L, Xu D, Zhang WJ, Tang Y, Peng Y. Astrocytes induce proliferation of oligodendrocyte progenitor cells via connexin 47-mediated activation of Chi3l1 expression. Eur Rev Med Pharmacol Sci. 2019;23(7):3012–20.

