## Supplemental Figures and Tables for "CHI3L1 (YKL-40) and Chit-1 expressing glia in the white matter of ALS, FTLD, and AD correlate to pathology and disease duration"

**A** **Original**

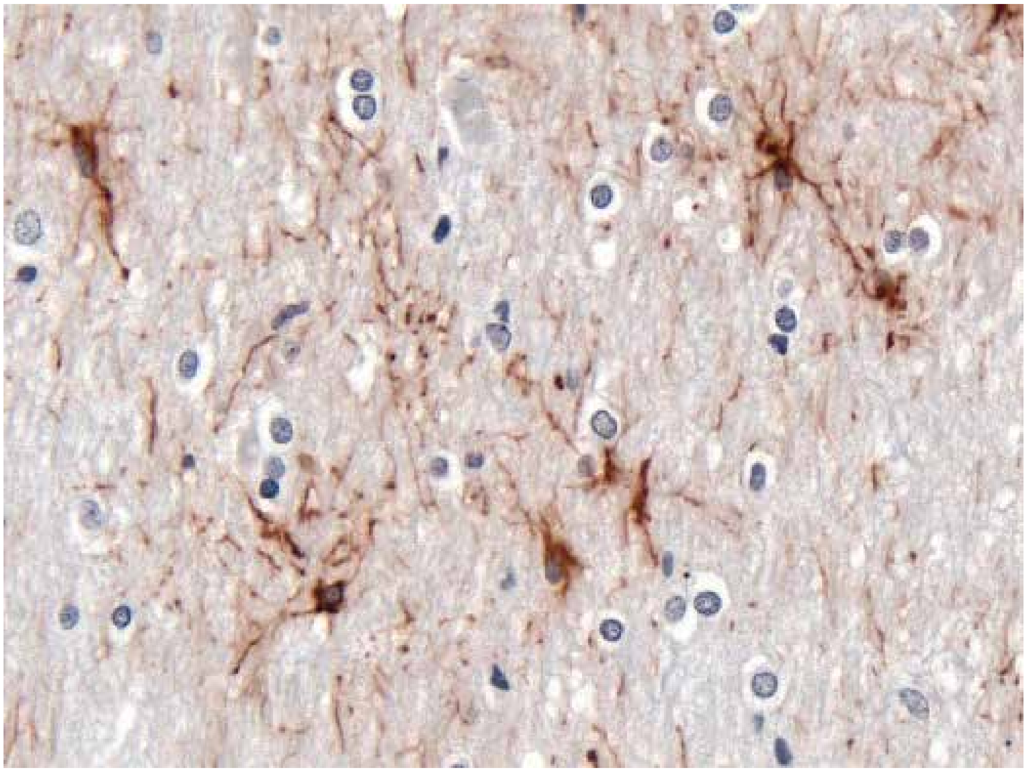

**B** **Masked**

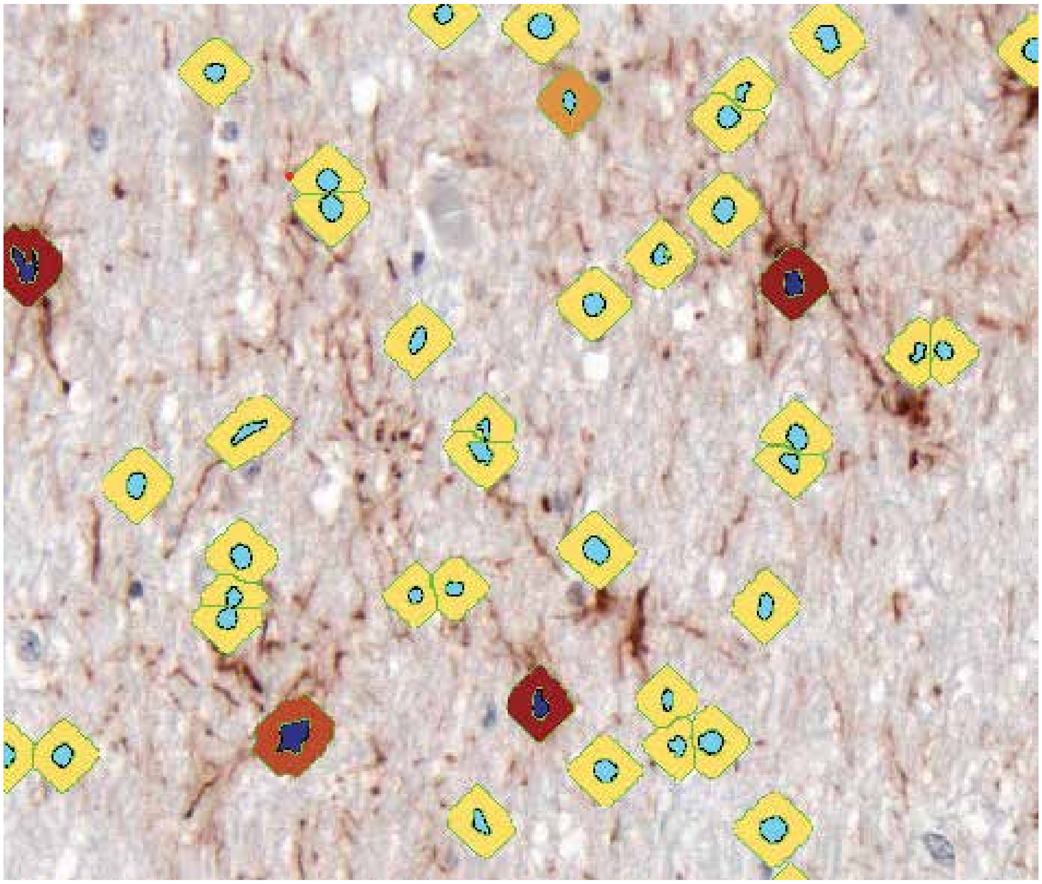

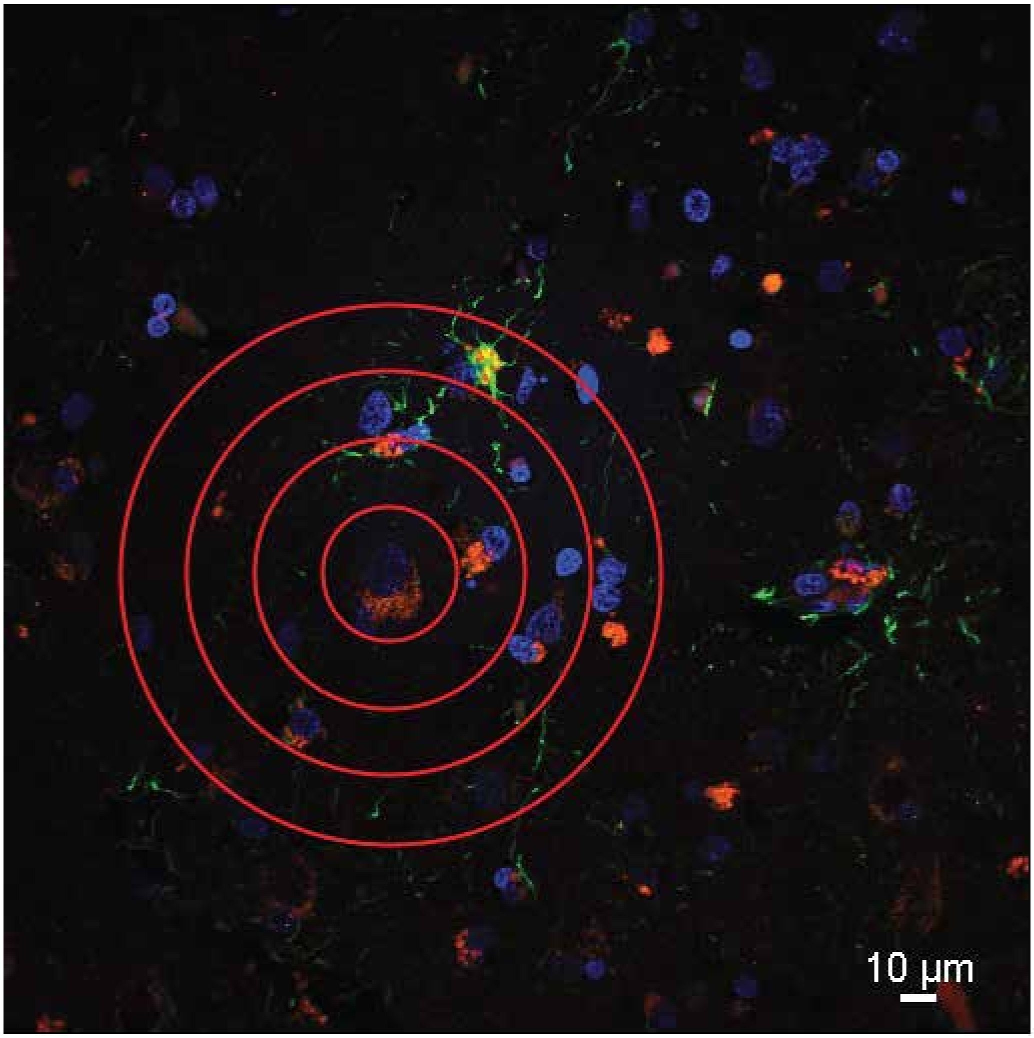
10 µm

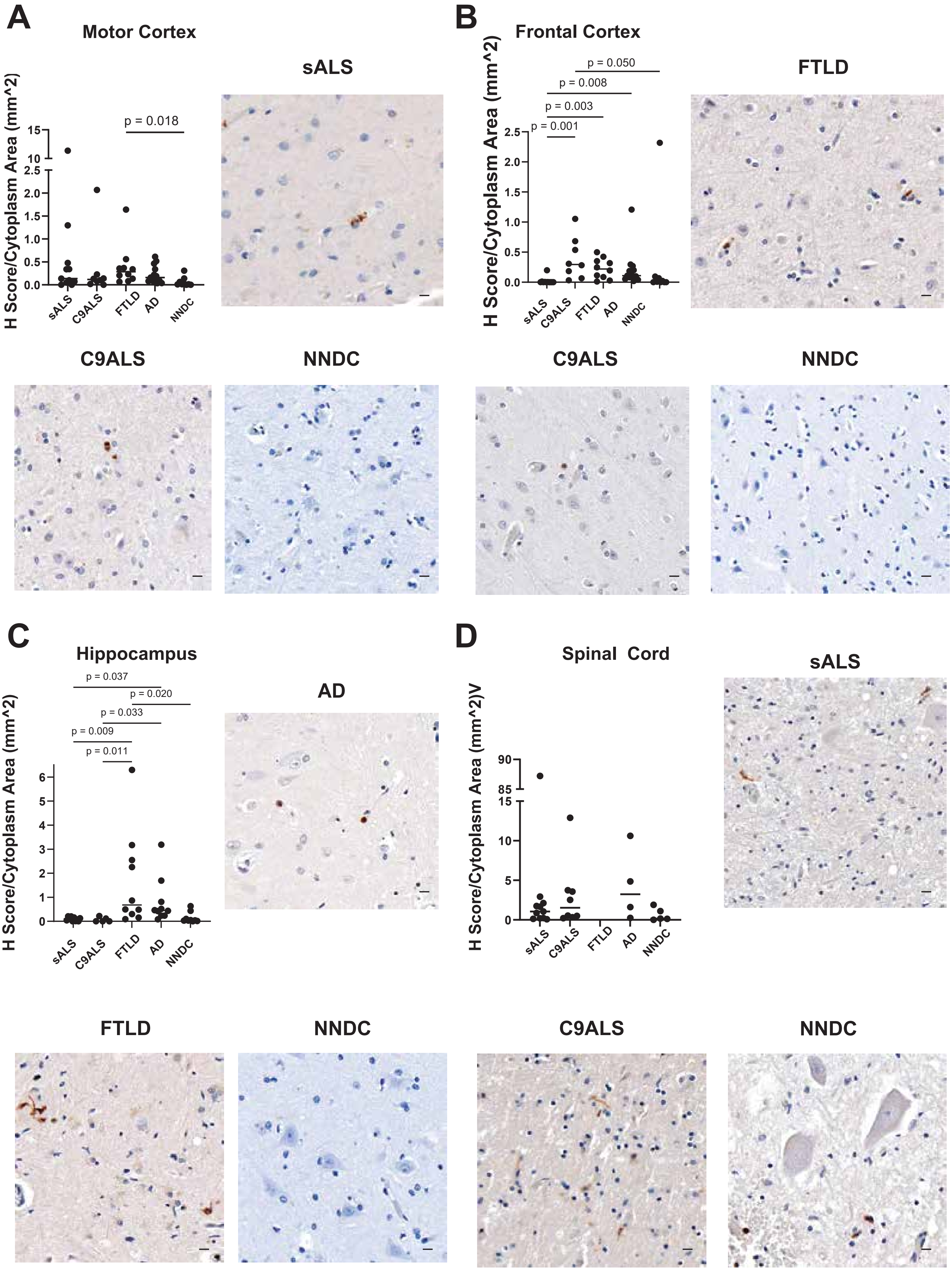
A
Motor Cortex
sALS
H Score/Cytoplasm Area (mm^2)
p = 0.018
15
10
2.5
2.0
1.5
1.0
0.5
0.0
sALS
C9ALS
FTLD
AD
NNDC
B
Frontal Cortex
FTLD
H Score/Cytoplasm Area (mm^2)
p = 0.050
p = 0.008
p = 0.003
p = 0.001
2.5
2.0
1.5
1.0
0.5
0.0
sALS
C9ALS
FTLD
AD
NNDC
C9ALS
NNDC
C9ALS
NNDC
C
Hippocampus
AD
H Score/Cytoplasm Area (mm^2)
p = 0.037
p = 0.020
p = 0.033
p = 0.009
p = 0.011
6
5
4
3
2
1
0
sALS
C9ALS
FTLD
AD
NNDC
D
Spinal Cord
sALS
H Score/Cytoplasm Area (mm^2)V
90
85
15
10
5
0
sALS
C9ALS
FTLD
AD
NNDC
FTLD
NNDC
C9ALS
NNDC

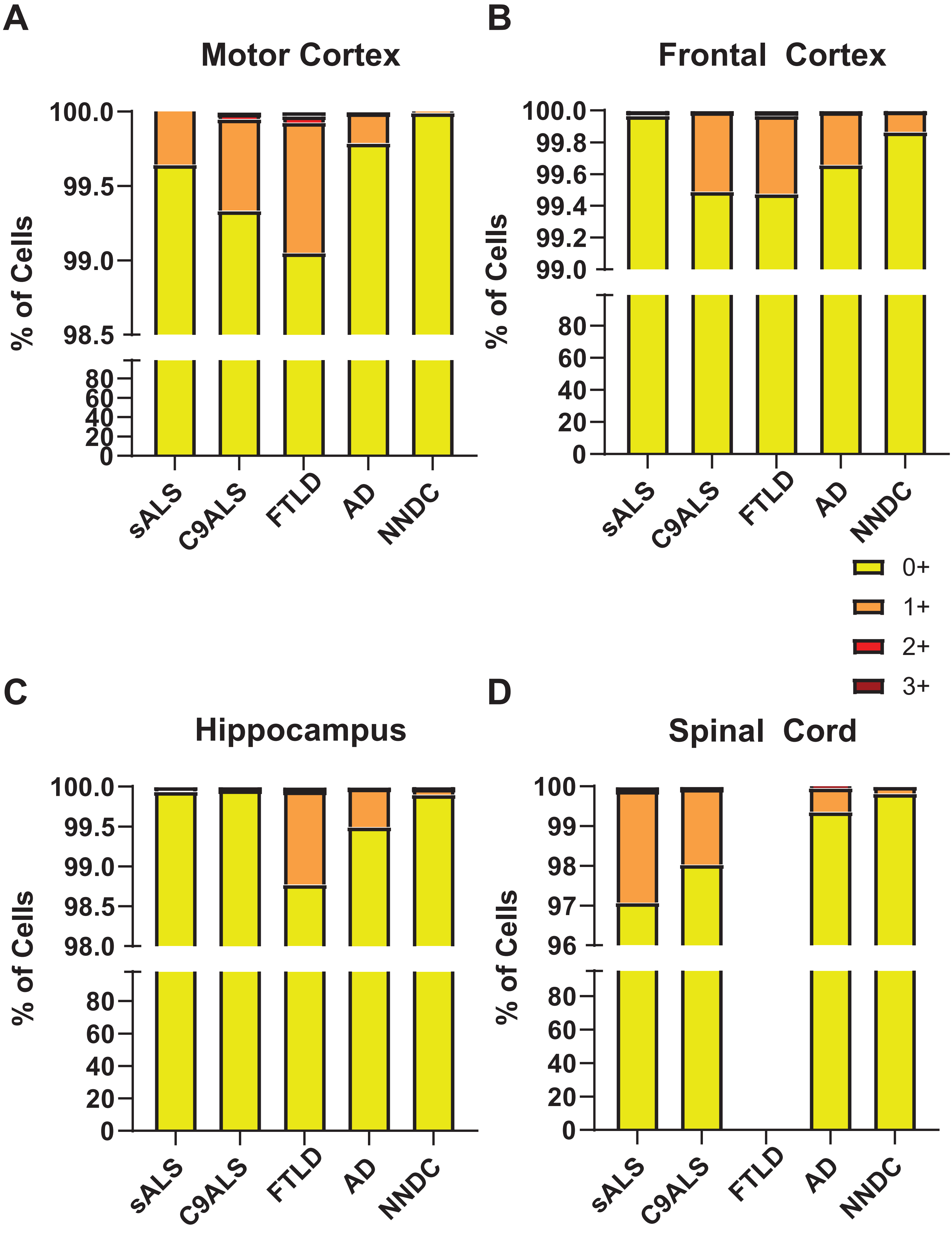

A
Motor Cortex
100.0
99.5
99.0
98.5
80
60
40
20
0
% of Cells
sALS
C9ALS
FTLD
AD
NNDC
B
Frontal Cortex
100.0
99.8
99.6
99.4
99.2
99.0
80
60
40
20
0
% of Cells
sALS
C9ALS
FTLD
AD
NNDC
0+
1+
2+
3+
C
Hippocampus
100.0
99.5
99.0
98.5
98.0
80
60
40
20
0
% of Cells
sALS
C9ALS
FTLD
AD
NNDC
D
Spinal Cord
100
99
98
97
96
80
60
40
20
0
% of Cells
sALS
C9ALS
FTLD
AD
NNDC

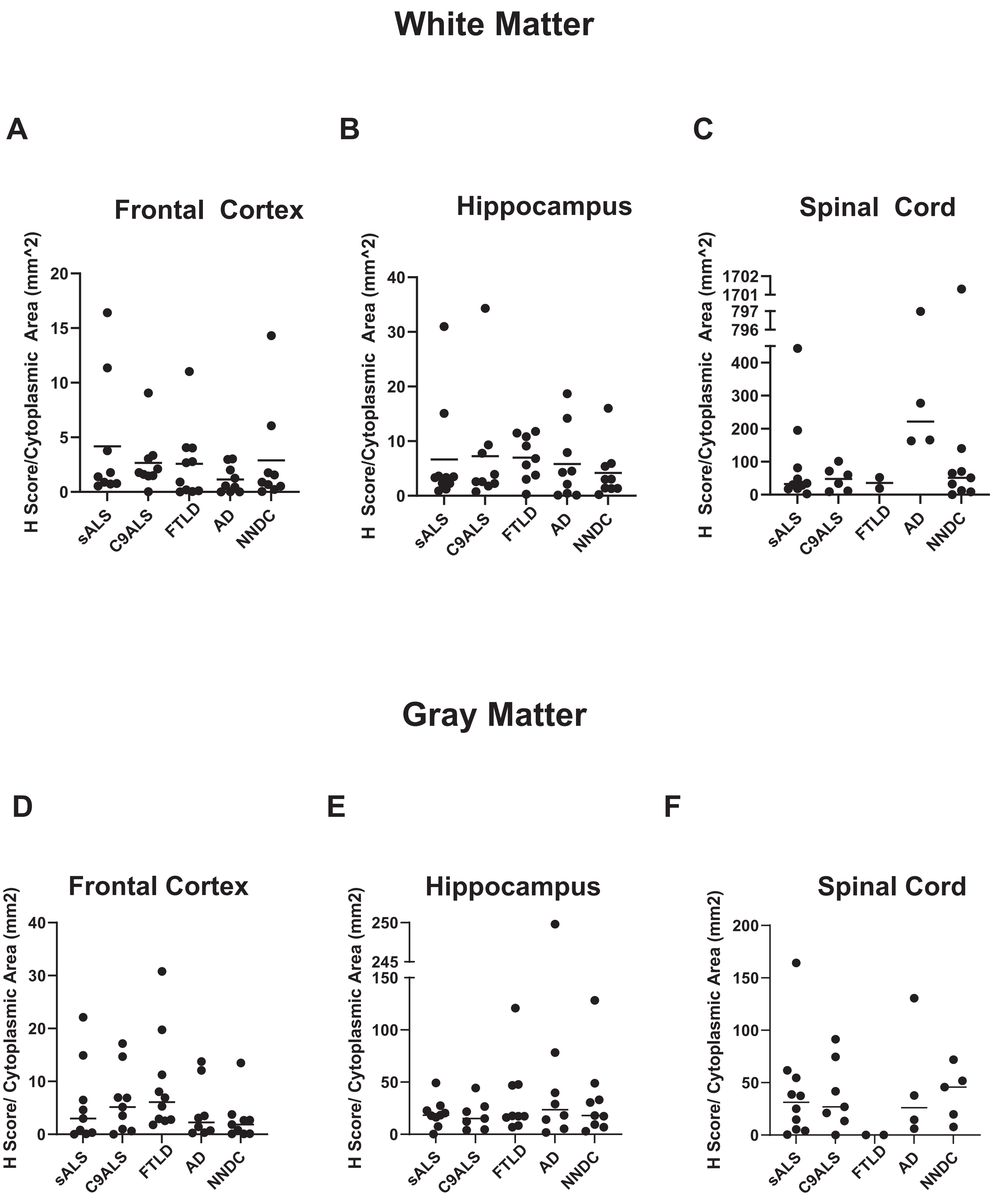
White Matter
A
Frontal Cortex
H Score/Cytoplasmic Area (mm^2)
20
15
10
5
0
sALS
C9ALS
FTLD
AD
NNDC
B
Hippocampus
H Score/Cytoplasmic Area (mm^2)
40
30
20
10
0
sALS
C9ALS
FTLD
AD
NNDC
C
Spinal Cord
H Score/Cytoplasmic Area (mm^2)
1702
1701
797
796
400
300
200
100
0
sALS
C9ALS
FTLD
AD
NNDC
Gray Matter
D
Frontal Cortex
H Score/ Cytoplasmic Area (mm2)
40
30
20
10
0
sALS
C9ALS
FTLD
AD
NNDC
E
Hippocampus
H Score/ Cytoplasmic Area (mm2)
250
245
150
100
50
0
sALS
C9ALS
FTLD
AD
NNDC
F
Spinal Cord
H Score/ Cytoplasmic Area (mm2)
200
150
100
50
0
sALS
C9ALS
FTLD
AD
NNDC

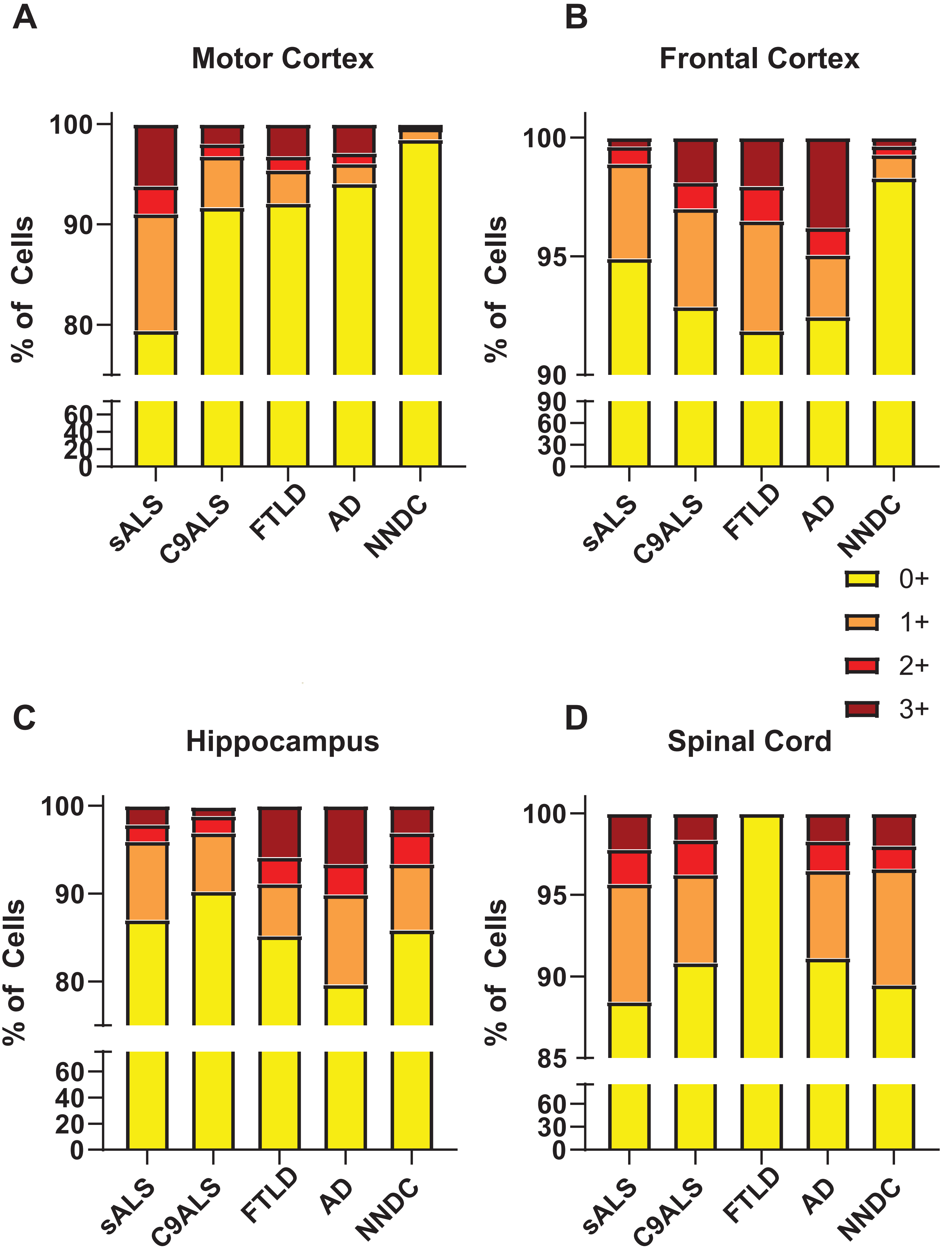
A
Motor Cortex
B
Frontal Cortex
C
Hippocampus
D
Spinal Cord
% of Cells
sALS
C9ALS
FTLD
AD
NNDC
0+
1+
2+
3+
100
90
80
95
85
60
40
30
20
0

| Antibody | Species | Dilution | Company | Catalog Number |
|---|---|---|---|---|
| Anti-CHI3L1 | Rabbit polyclonal | 1:300 | ThermoFisher | PA5-43736 |
| Anti-Chit-1 | Goat polyclonal | 2ug/mL | R&D Systems | AF3559 |
| Anti-TDP43 Phospho | Rat monoclonal | 1:500 | Biolegend | SIG-39852 |
| Biotinylated Goat anti-Rabbit IgG | Anti-Rabbit | 1:200 (IHC) | Vector Labs | BA-1000 |
| Biotinylated Rabbit anti-Goat IgG | Anti-Goat | 1:200 (IHC) | Vector Labs | BA-5000 |
| Alexa Fluor 488 Donkey anti-Goat IgG | Anti-Goat | 1:200 (IF) | Invitrogen | A-11055 |
| Alexa Fluor 488 Goat anti-Rabbit IgG | Anti-Rabbit | 1:200 (IF) | Invitrogen | A-11008 |
| Alexa Fluor 555 Goat anti-Rat IgG | Anti-Rat | 1:200 (IF) | Invitrogen | A-21434 |
| Alexa Fluor 647 Goat anti-Rabbit IgG | Anti-Rabbit | 1:200 (IF) | Invitrogen | A-21244 |

Supplemental Figure Legends

Figure S1: Aperio ImageScope Cytoplasmic Algorithm applied to the (A) original image. (B) Masked image with three intervals of staining intensity. Nuclei (blue), negative cytoplasmic staining (yellow), weak positive staining (orange), moderate positive staining (red-orange), strong positive staining (burgundy). This image was taken at 20x magnification.

Figure S2: Concentric Circles Plugin on ImageJ Software. Circles were centered on a nucleus of a neuron with or without pathological TDP and used to measure the distance from Chit-1+ or CHI3L1+ cells from the neuron. The distance between each circle is 20μm.

Figure S3: Immunohistochemistry for Chit-1 in the gray matter of patients with sALS, C9ALS, FTLD, and NNDC. Representative images used for quantification of Chit-1-positive cells in the gray matter of the (A) motor cortex, (B) frontal cortex, (C) hippocampus, and (D) lumbar spinal cord. Data analysis was performed in a blinded manner as described in Methods. Pairwise comparisons by Kruskal-Wallis test with Dunn post-hoc correction were used to assess differences between subject groups for each cortical and spinal cord region. *P* values <0.05 shown. Chit-1, chitotriosidase-1; sALS, sporadic amyotrophic lateral sclerosis; C9ALS, C9orf72 ALS; FTLD, frontotemporal lobar degeneration; NNDC, non-neurologic disease control. All images at 20X magnification and scale bar = 10 μm.

Figure S4: Percentage of cells expressing Chit-1 for each defined staining intensity (0+, 1+, 2+, 3+) in the gray matter of the (A) motor cortex, (B) frontal cortex, (C) hippocampus, and (D) lumbar spinal cord of sALS, C9ALS, FTD, AD, and NNDC patients. 0+: no expression (yellow), 1+: weak positive expression (orange), 2+: moderate positive expression (red), 3+: strong positive expression (burgundy). Chit-1, chitotriosidase-1; sALS, sporadic amyotrophic lateral sclerosis; C9ALS, C9orf72 ALS; FTLD, frontotemporal lobar degeneration; NNDC, non-neurologic disease control.

Figure S5: Quantification of CHI3L1-positive cells in the white matter of (A) frontal cortex, (B) hippocampus, and (C) spinal cord as well as the gray matter of (D) frontal cortex, (E) hippocampus, and (F) spinal cord, using 5 images from each case. Data analysis was performed in a blinded manner and the average value for each case plotted on the graphs. A Kruskal-Wallis test was used to assess differences between pairwise comparisons, and *p* values were corrected using the Dunn post-hoc correction. *P* values <0.05 shown. sALS, sporadic amyotrophic lateral sclerosis; C9ALS, C9orf72 ALS; FTLD, frontotemporal lobar degeneration; NNDC, non-neurologic disease control.

Figure S6: Percentage of cells expressing CHI3L1 for each defined staining intensity (0+, 1+, 2+, 3+) in the gray matter of the (A) motor cortex, (B) frontal cortex, (C) hippocampus, and (D) lumbar spinal cord of sALS, C9ALS, FTD, AD, and NNDC patients. 0+: no expression (yellow), 1+: weak positive expression (orange), 2+: moderate positive expression (red), 3+: strong positive expression (burgundy). CHI3L1, Chitinase-3-like 1; sALS, sporadic amyotrophic lateral sclerosis; C9ALS, C9orf72 ALS; FTLD, frontotemporal lobar degeneration; NNDC, non-neurologic disease control.

Table S1: Antibody List
